# Unraveling Shared Pathways: A Comprehensive Systematic Review of Common Fiber Tracts in Amyotrophic Lateral Sclerosis and Frontotemporal Dementia using Diffusion Tensor Imaging

**DOI:** 10.1101/2023.11.09.23298318

**Authors:** Ashwag Rafea S Alruwaili, Matthew Devine, Pamela Mccombe

## Abstract

This systematic review evaluated MRI studies of fibre tract abnormalities in patients with amyotrophic lateral sclerosis (ALS) and/or fronto-temporal dementia (FTD). After searching 5 databases, 63 papers met inclusion criteria reporting 1674 patients and 1411 healthy controls. The papers studied a range of fibre tracts. Techniques used included overall comparisons and regions of interest. All papers reported results of fractional anisotropy (FA) and some also reported other DTI metrics. In ALS, the hallmark feature of cortico-spinal tracts (CST) involvement is consistently found, while in FTD the only part of the motor tracts that was found to show changes is the corona radiata (CR). The review also highlighted overlapping abnormalities between ALS and FTD, suggesting that these conditions exist on a spectrum. Both ALS and FTD exhibited CST abnormalities, with extra-motor involvement in the cingulum and the CC. Many tracts including the corpus callosum (CC) and cingulum (Cg), the superior longitudinal fasciculus (SLF) and inferior fronto-occiptal fasciculus (IFOF), were abnormal in both ALS and FTD. The integrity of specific white matter tracts, such as the uncinated fasciculus (uncF), forceps minor, and callosal radiation, appeared critical for cognitive functions related to Theory of Mind, cognitive control, and emotion recognition.There was, however, extramotor involvement in ALS.

## 3 Introduction

Amyotrophic lateral sclerosis (ALS) is a fatal neurodegenerative disease characterized by the progressive death of upper and lower motor neurons. ALS is thought to arise from a combination of genetic susceptibility and environmental exposure (Fang, Kamel et al. 2009, Sutedja, Veldink et al. 2009). Patients with ALS develop weakness of the limbs, the bulbar muscles and the diaphragm (Kiernan, Vucic et al. 2011). There are clinical signs of dysfunction of both upper motor neurons (UMN) and lower motor neurons (LMN) (Miller, Gelinas et al. 2005).

Frontotemporal dementia (FTD) is the term for progressive dementias characterized by cognitive and behavioral difficulties, with degeneration of the frontal and temporal lobes. FTD can be divided into behavioral variant, primary progressive aphasia and semantic dementia (Warren, Rohrer et al. 2013). Pathological studies allow the division of FTD into tauopathy and ubiquinopathy (Kertesz and McMonagle 2011).

A proportion of patients with ALS show cognitive impairment with subtle executive function deficits (Raaphorst, De Visser et al. 2010, Phukan, Elamin et al. 2012) and 10- 15% fulfill the criteria for diagnosis of frontotemporal dementia (FTD) (Ringholz, Appel et al. 2005). It is now considered that ALS and FTD lie on a pathological and clinical spectrum (Von Braunmühl 1932). Some causative genes are found in both ALS and FTD (Siddique and Deng 1996, Al-Chalabi, Jones et al. 2012, Pan and Chen 2013), with the most important so far identified being C9orf72 (Mori, Weng et al. 2013). Some pathological features are common to FTD, ALS-FTD and ALS (Ferrari, Kapogiannis et al. 2011), particularly TDP-43 pathology and possibly dipeptide repeats (Al-Chalabi and Hardiman 2013), which are found in many patients with ALS and patients with FTD with ubiquinopathy. An anatomical study has shown that behavioural disturbance in patients with ALS is significantly associated with frontal lobe involvement (Tsujimoto, Senda et al. 2011).

Pathological studies on autopsy material are done at the end stage of disease, whereas imaging studies allow the assessment of earlier stages of disease. Various neuroimaging modalities have been used to study ALS and FTD (Wang, Poptani et al. 2006) including diffusion tensor imaging (DTI), which is able to detect microstructural changes in fiber tracts(Uluğ, Moore et al. 1999, Hong, Lee et al. 2004). DTI is increasingly being used to show changes in fiber tracts in neurodegenerative diseases. Generally speaking, decreased diffusivity and increased diffusion anisotropy reflect greater structural organization and white matter (WM) structural integrity (Pannek, Guzzetta et al. 2012).

There have been previous meta-analyses of DTI abnormalities in ALS. One study reported a voxel based meta-analysis of FA of 143 non-demented ALS patients from 7 studies (Li, Pan et al. 2012). Another reported 170 patients from 9 studies, to determine a threshold for FA abnormality in the corticospinal tract (Rosskopf, Müller et al. 2015).

Because some patients with ALS have FTD or less severe cognitive defects, we expect there will be overlap of pathology (Yamada, Sakai et al. 2009). Brain regions that show abnormal DTI changes in both ALS and FTD may provide clues to elucidating the pathogenesis of ALS-FTD overlap. Here I aim to review the literature regarding the location and extent of white matter tract degeneration in ALS patients with and without FTD, and patients with pure FTD, to determine if there is overlap of the fiber tract involvement in these conditions. The studies in this review use various MRI measures making a meta-analysis impossible. This systematic review is a therefore a descriptive study. We also review the correlations of DTI measures with clinical status.

## 4 Methods

### 4.1 Search strategy and key words

In December 2015, a systematic search was conducted of the databases; Pubmed Embase, CINAHL, Scopus and Web of Science.

The search strategy used the medical subject heading (MeSH) terms and the keywords: “amyotrophic lateral sclerosis”, “ALS”, “frontotemporal dementia”, “FTD”, “cognitive impairment”, “diffusion tensor imaging”, “DTI”, “tractography”, and “white matter tracts”. These were then combined to limit the findings: [“amyotrophic lateral sclerosis” AND diffusion tensor imaging], [“Frontotemporal dementia” AND diffusion tensor imaging], [amyotrophic lateral sclerosis” AND tractography], [“Frontotemporal dementia” AND tractography], [“Frontotemporal dementia” AND white matter tracts], [amyotrophic lateral sclerosis” AND white matter tracts].

### 4.2 Selection criteria

#### Inclusion criteria

1) studies were carried out in human subjects equal to or older than 18 years of age; 2) subjects diagnosed with amyotrophic lateral sclerosis met the accepted El Escorial diagnostic criteria (Brooks, Miller et al. 2000); 3) subjects diagnosed with ALS-FTD or FTD fulfilled the Neary criteria for diagnosis of frontotemporal dementia (Neary, Snowden et al. 1998); 4) studies provided an adequate descriptions of the patient sample such as male: female ratio, age and whether the subjects had other neurological disorders or disease; 5) studies used diffusion tensor imaging (DTI) region of interest (ROI), tractography, or voxel-based analysis to assess white matter tracts by comparing DTI measures between patients and healthy controls; 6) studies provided adequate description of the imaging protocol (MR sequence, b-value, diffusion directions, threshold) and statistical data such as mean, and SD; 7) studies measured DTI metrics in specific white matter regions with defined coordinates.

#### Exclusion criteria

1) studies which included children and adolescents younger than 18 years; 2) subjects who had associated neurological disorders or pathologic changes such as brain tumor, epilepsy or brain injury; 3) studies with fewer than 7 participants, 4) studies without healthy controls participants; 5) studies not published in English. 6) studies with missing data such as details about participants or imaging sequence. 7) studies assessed as low quality studies.

Two independent raters (A.A and M.D) assessed eligibility based on the title and abstract of each study. Then a full screening of references was performed for inclusion.

### 4.3 Assessment of methodological quality

Two raters (Ashwag Alruwaili, Mathew Devine) independently assessed the quality of each study according to the quality of diagnostic accuracy studies (QUADAS) criteria (Whiting, Rutjes et al. 2003). Each criterion was scored “yes”, “no” or “unclear”. A score of 7-10 of 14 is considered to be a high quality study.

### 4.4 Data extraction

The details of data extracted from all eligible studies included: a) total number of participants, 2) the imaging sequences (field strength, b-value, number of directions, smoothing and FA threshold), 3) WM analysis type (ROI, VBA, tractography), 4) DTI metrics (FA, MD, AD, RD) and, 5) the brain regions studied.

### 4.5 Selection of tracts for inclusion in figures

To create a visual representation of the tracts that are abnormal in ALS and FTD, we selected tracts that were reported as being significantly abnormal in more than 66% of the studies that assessed them. These are the tracts for which the evidence is strongest. Selection of these tracts was based on results of FA, because this measure was used most consistently across the different studies.

## 5 Results

### 5.1 Overall summary and participant characteristics

As shown in Figure 4.1, of 233 papers, a total of 64 studies met the criteria for full review. These papers reported on a total 1674 patients (ALS N=1192, FTD N=458 and ALS-FTD N=24) and 1411 healthy controls. The extracted data are given in Supp Table 4.1. Table 4_1 summarizes the information about the number of participants in each clinical category. For the studies of ALS, the total patient number was 1192, with 991 controls. For the studies of FTD there were 458 subjects and 420 controls. In addition, two paper studied ALS with FTD (n=24).

**Figure 4.1:**
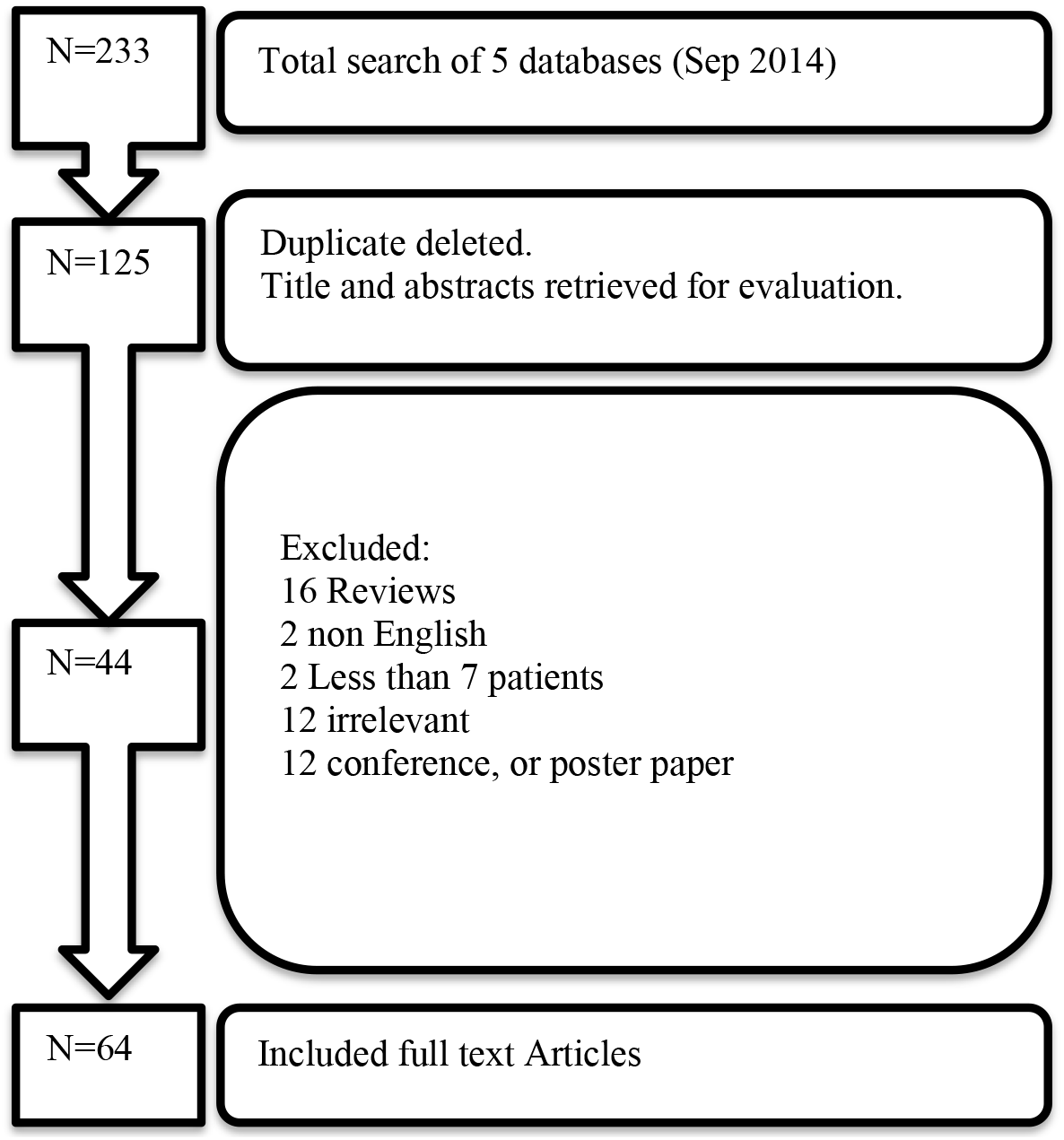
Pipeline of inclusion of studies.

**Table 4.1:** Details of participants.

| Disease studied | No of papers | No of patients | No of controls |
| --- | --- | --- | --- |
| **ALS** | 43 | 1192 | 991 |
| **FTD** | 20** | 458 | 420 |
| **ALS and FTD** | 2 | 24 | * |

\* Controls included in the main ALS studies controls column.
\*\* Three ALS papers have included ALS-FTD subjects (so total of 23 studies).

Table 4_2 summarizes the use of different techniques to analyze white matter changes in ALS, FTD and ALS-FTD. These were regions of interest (ROI, n=23), tractography (n=15), tract-based spatial statistics (TBSS, n=22), and tract of interest (TOI, n=2). There were also studies that used mixed analysis (n=14).

**Table 4.2:** Summary of WM analysis.

| Disease studied | No. of studies using each technique |  |  |  |  |
| --- | --- | --- | --- | --- | --- |
|  | ROI | Tractography | TBSS | TOI | Mixed |
| **ALS** | 16 | 12 | 15 | 1 | 11* |
| **FTD** | 7 | 3 | 6 | 1 | 3** |
| **ALS and FTD** | - | - | 1 | - | - |
| *1 ROI+TBSS+ Tractography, 3 ROI+TBSS, 3 TBSS+Tractography, 1 ROI + Tractography, 1TOI+ROI |  |  |  |  |  |
| ** 1 TOI+VWA, 1 canonical correlation analysis, 1 ROI+TBSS |  |  |  |  |  |

**Table 4.3:**
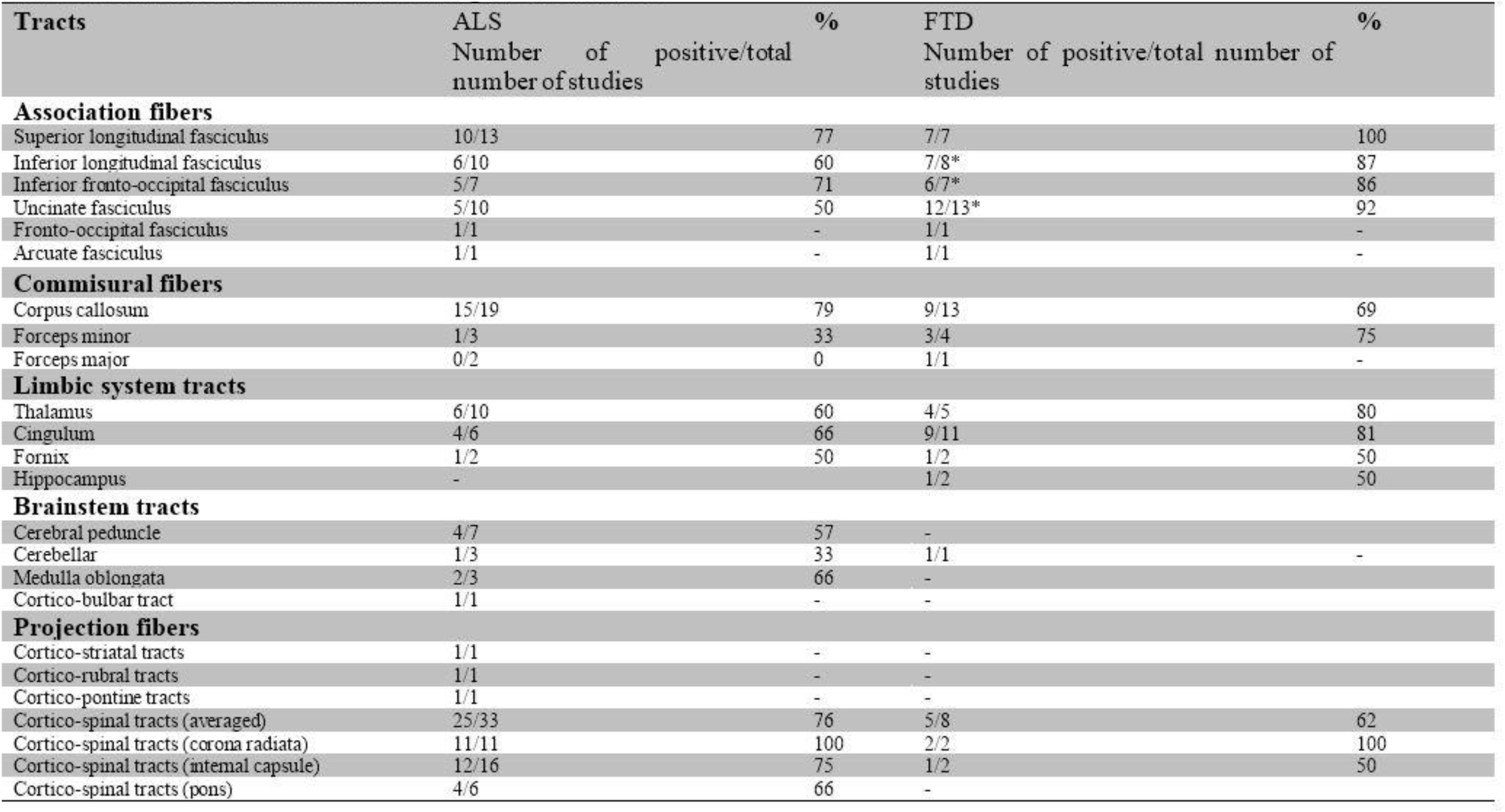
Numbers of FA stuclies with significant results.

#### 5.1.1 DTI in ALS

A summary of the number of papers that studied each tract, and the number of these papers that found significant abnormalities in each tract using FA is provided in Table 4.3. The full details of the extracted data are presented in Supp Table 4_1. In the following sections, we mostly describe the studies that found significant results from DTI measures. The non-significant findings are presented in each table but not within the text. The full details of the studies with both positive and negative results, for each class of white matter fibers, are presented in Tables 4.4–4.7. A visual summary of the WM tracts that were found to be significant in ALS studies is presented in Figure 4.2. The details of the clinical correlations of the DTI results for each MRI metric are given in Supp Tables (4.2–4.6).

##### 5.1.1.1 Whole brain analysis

Six studies performed whole brain analysis (Lillo, Mioshi et al. 2012, McMillan, Brun et al. 2012, Pettit, Bastin et al. 2012, Prudlo, Bissbort et al. 2012, Sarica, Cerasa et al. 2014, Trojsi, Caiazzo et al. 2015). Whole brain TBSS found significant changes in FA and MD in the SLF bilaterally (Sarica, Cerasa et al. 2014, Trojsi, Caiazzo et al. 2015). Based on the clinical staging system for ALS, proposed by Roche et al. (Roche, Rojas-Garcia et al. 2012) (refer to appendix for details), a longitudinal study revealed that the body of CC shows reduced FA in stage 2A, and reduced FA and increased MD and RD in stage 3, compared to controls (Trojsi, Caiazzo et al. 2015). A different study found that the FA in the SLF, ILF and IFOF was reduced in ALS patients compared to controls (Prudlo, Bissbort et al. 2012). In both bulbar and limb onset ALS compared to controls, CST showed reduced FA along the tract (Cardenas-Blanco, Machts et al. 2014). One study reported that ALS patients showed more degeneration in the CST than in callosal and association tracts compared to controls (Lillo, Mioshi et al. 2012). While this was not the case when the same ALS subjects were compared to FTD and ALS-FTD patients, TBSS results shows less CST involvement than extra-motor tracts (Lillo, Mioshi et al. 2012).

##### 5.1.1.2 ROI and Tractography analysis

###### 5.1.1.2.1 Projection Fibers

Corticospinal tract (CST) degeneration is a hallmark of ALS and hence the CST was studied in ALS frequently. The studies that found significant and non-significant results are given in Table 4_4 and a summary in Figure 4.2.

**Table 4.4:**
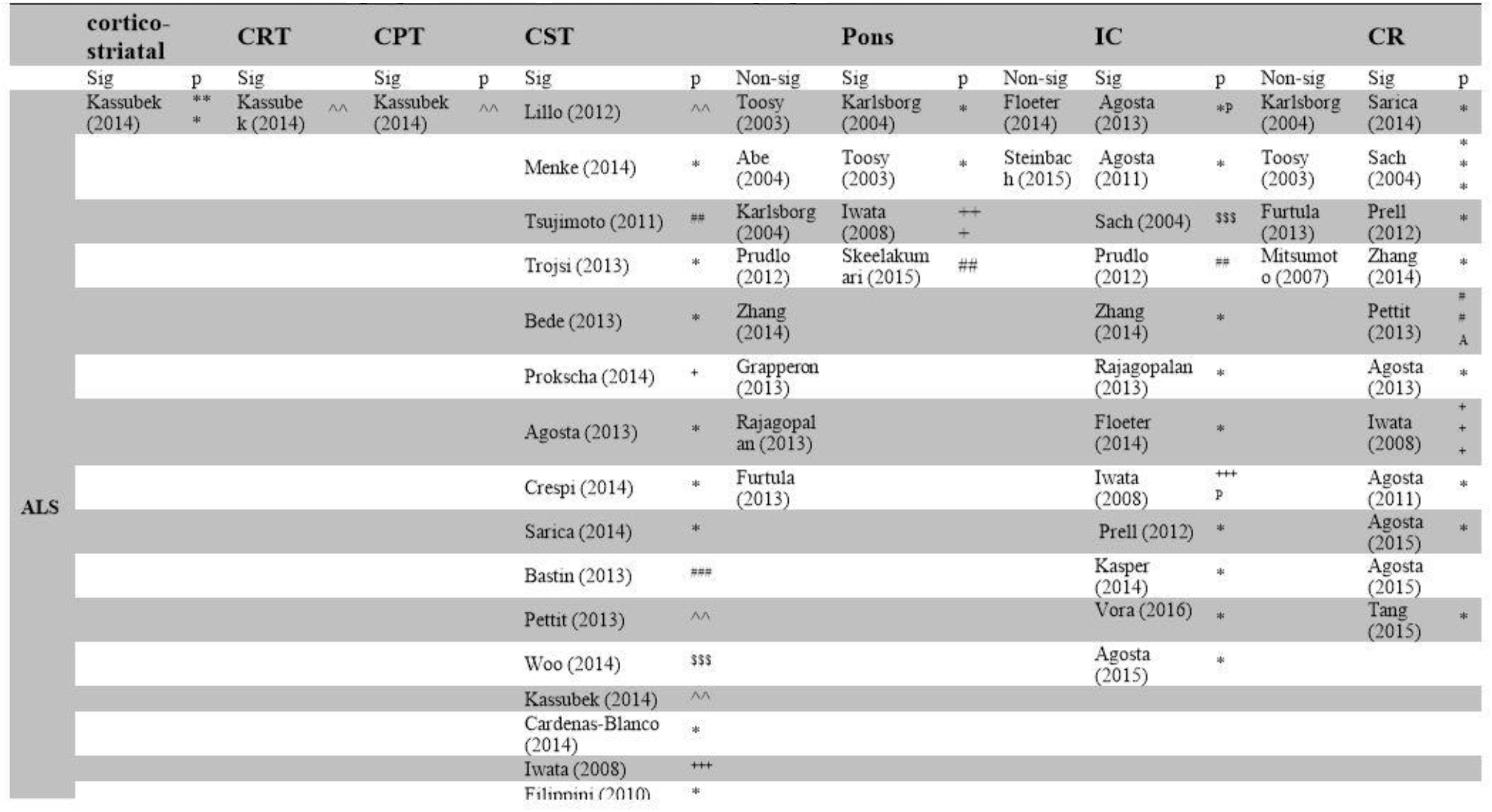
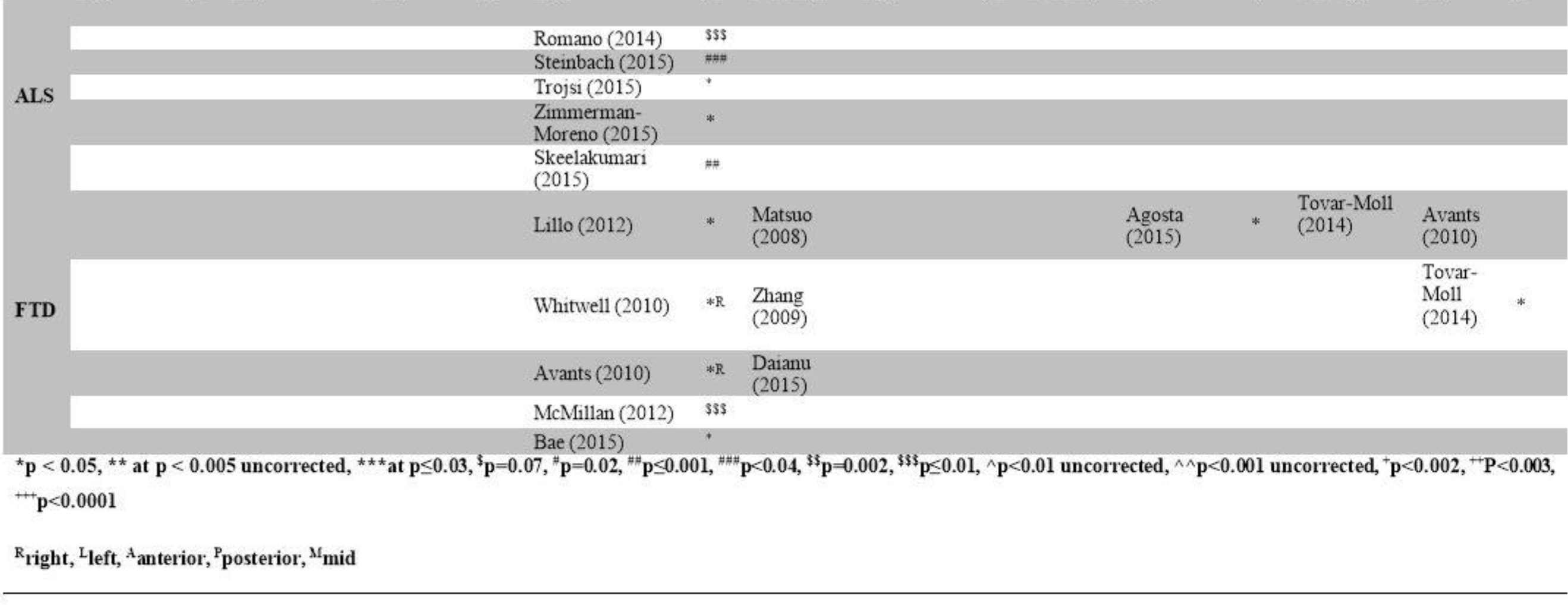
Stuclies of FA iu the rojection fibers, which stucliecl the rojection fibers.

Apart from the study of Grapperon et al who reported an increase in FA (Grapperon, Verschueren et al. 2014), all other studies found reduction of the average FA over the entire CST(Abe, Yamada et al. 2004, Karlsborg, Rosenbaum et al. 2004, Iwata, Aoki et al. 2008, Sarro, Agosta et al. 2011, Tsujimoto, Senda et al. 2011, Lillo, Mioshi et al. 2012, Pettit, Bastin et al. 2012, Prudlo, Bissbort et al. 2012, Agosta, Caso et al. 2013, Bastin, Pettit et al. 2013, Rajagopalan, Yue et al. 2013, Trojsi, Corbo et al. 2013, Crespi, Cerami et al. 2014, Floeter, Katipally et al. 2014, Kassubek, Müller et al. 2014, Prokscha, Guo et al. 2014) as well as at different levels along the tract (Toosy, Werring et al. 2003, Karlsborg, Rosenbaum et al. 2004, Mitsumoto, Ulug et al. 2007, Rajagopalan, Yue et al. 2013, Hübers, Müller et al. 2015). This was more apparent in the right CST (Sarica, Cerasa et al. 2014) with RD increase (Zhang, Schuff et al. 2009), superiorly (Bede, Bokde et al. 2013) caudally (Zhang, Yin et al. 2014, Hübers, Müller et al. 2015) and rostrally (Filippini, Douaud et al. 2010) and with MD increases (Toosy, Werring et al. 2003, Prudlo, Bissbort et al. 2012, Floeter, Katipally et al. 2014) at the level of the internal capsule (Sach, Winkler et al. 2004). One study found changes in CST in familial ALS (fALS) but not in sporadic ALS when compared to controls (Mitsumoto, Ulug et al. 2007). In both bulbar and limb onset ALS compared to controls, CST showed reduced FA and exhibited in higher RD and MD when using ROI (Cardenas-Blanco, Machts et al. 2014). Some other groups reported significant changes in RD (Menke, Körner et al. 2014, Zimmerman-Moreno, Ben Bashat et al. 2015, Vora, Kumar et al. 2016) and AD (Menke, Körner et al. 2014) along the CST (Kassubek, Müller et al. 2014, Menke, Körner et al. 2014). Steinbach et al found connectivity indices^1^ to be significantly reduced along the tract (Steinbach, Loewe et al. 2015). The motor pathway was also studied at the level of internal capsule (PLIC) (Kasper, Schuster et al. 2014, Vora, Kumar et al. 2016) and corona radiata where it was found to have significantly reduced FA (Agosta, Caso et al. 2013, Agosta, Galantucci et al. 2015) and increased RD (Sarica, Cerasa et al. 2014) and MD (Pettit, Bastin et al. 2012, Sarica, Cerasa et al. 2014, Agosta, Galantucci et al. 2015). Some studies investigated the CST at the level of pons (Toosy, Werring et al. 2003, Iwata, Aoki et al. 2008, Sheelakumari, Madhusoodanan et al. 2015) and found significantly reduced FA and increased MD. In the CST, Romano et al found significantly reduced FA and increased MD and RD, but no changes in AD (Romano, Guo et al. 2014). Another group classified ALS participants based on a staging system (Roche, Rojas-Garcia et al. 2012) and found reduced FA in left rostral CST in stages 2A; at diagnosis, and 3; when a third body region was involved (Trojsi, Caiazzo et al. 2015). The changes in the latter stage involved more DTI metrics such as higher MD and RD (Trojsi, Caiazzo et al. 2015).

The correlations of DTI measures with quantitative clinical measures are shown in Supp Tables 4.2–4.6. In ALS, increased AD (Menke, Körner et al. 2014), MD (Menke, Körner et al. 2014) and RD (Menke, Körner et al. 2014) and decreased FA along the CST were associated with higher UMN scores (Abe, Yamada et al. 2004, Iwata, Aoki et al. 2008, Filippini, Douaud et al. 2010, Trojsi, Corbo et al. 2013, Menke, Körner et al. 2014, Woo, Wang et al. 2014). Longer disease duration correlated with increased FA (Filippini, Douaud et al. 2010) and AD (Rajagopalan, Yue et al. 2013). Reduced FA along the CST correlated positively with delayed central motor conduction time (CMCT) and cortical-brainstem conduction time (CTX-BS) (Iwata, Aoki et al. 2008), but negatively with disease progression (Bastin, Pettit et al. 2013). Disease progression correlated with increased RD when using ROI (Menke, Körner et al. 2014) and correlated negatively with reduced connectivity indices (Steinbach, Loewe et al. 2015). In many studies (Furtula, Johnsen et al. 2013, Grapperon, Verschueren et al. 2014, Woo, Wang et al. 2014) there was no significant correlation between DTI metrics and clinical features such as forced vital capacity (FVC) and site of onset (Pettit, Bastin et al. 2012). Changes in FA (Woo, Wang et al. 2014), RD and MD were negatively correlated with ALSFRS-R (Sarica, Cerasa et al. 2014) in the right CST. The latter study found that there was a trend towards significant correlation between ALSFRS-R scores and FA (Sarica, Cerasa et al. 2014). Other studies found no correlation between FA and ALSFRS-R and disease duration (Furtula, Johnsen et al. 2013, Woo, Wang et al. 2014) or disease progression (Trojsi, Corbo et al. 2013).

When using cognitive examination, it was found that reduced FA correlated with reduced letter fluency (Pettit, Bastin et al. 2012) and negatively with frontal system behavior scale (FrSBe) score at the level of pontomesencephalic junction (Trojsi, Corbo et al. 2013). Reduced FA along CST correlated with Trail-making test scores (Sarro, Agosta et al. 2011) and Stroop test scores (Sarro, Agosta et al. 2011).

###### 5.1.1.2.2 Association Fibers

Studies finding both significant and non-significant results for each of the association fiber tracts in ALS are presented in Table 4_5 and Figure 4.2.

**Table 4.5:**
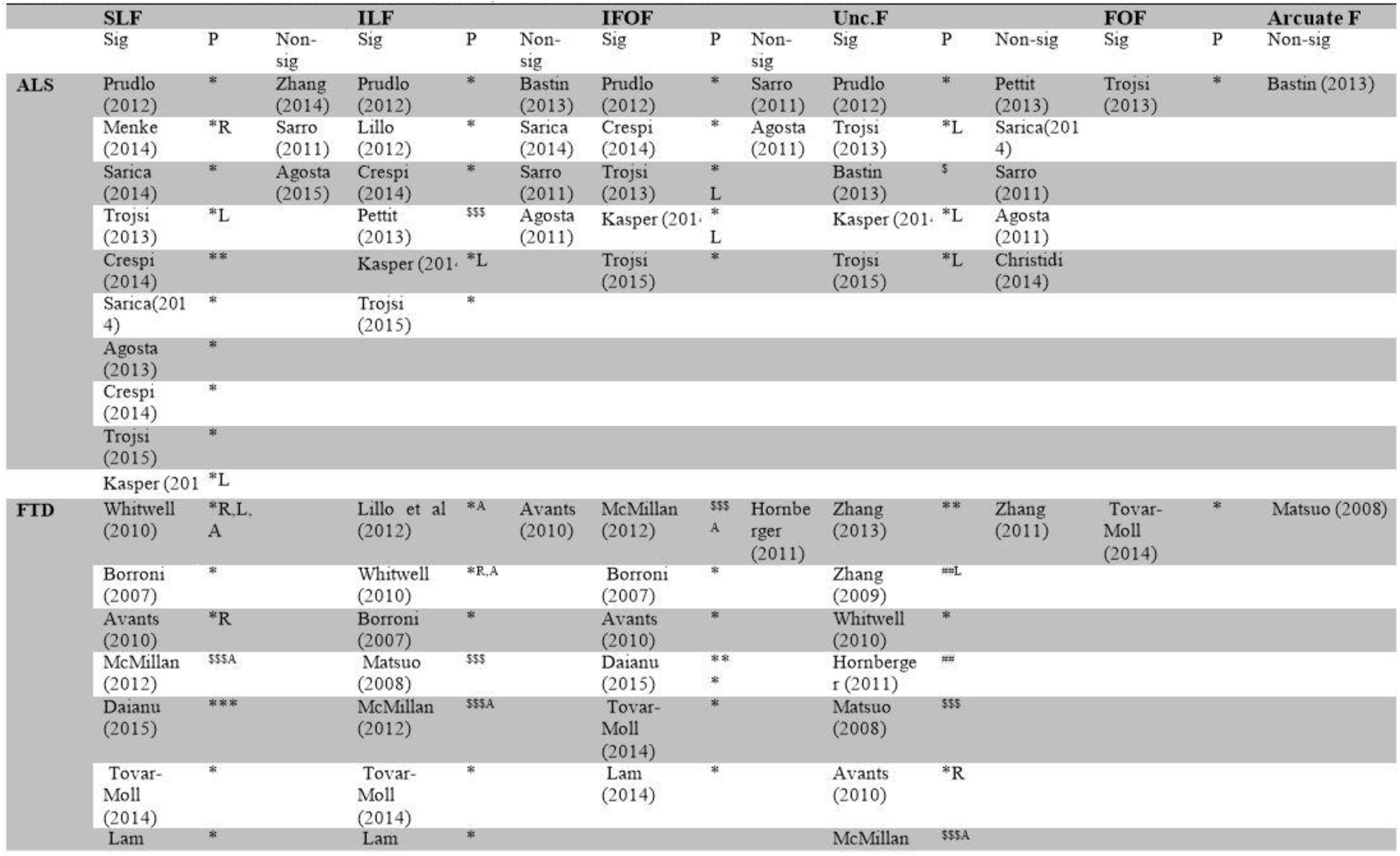
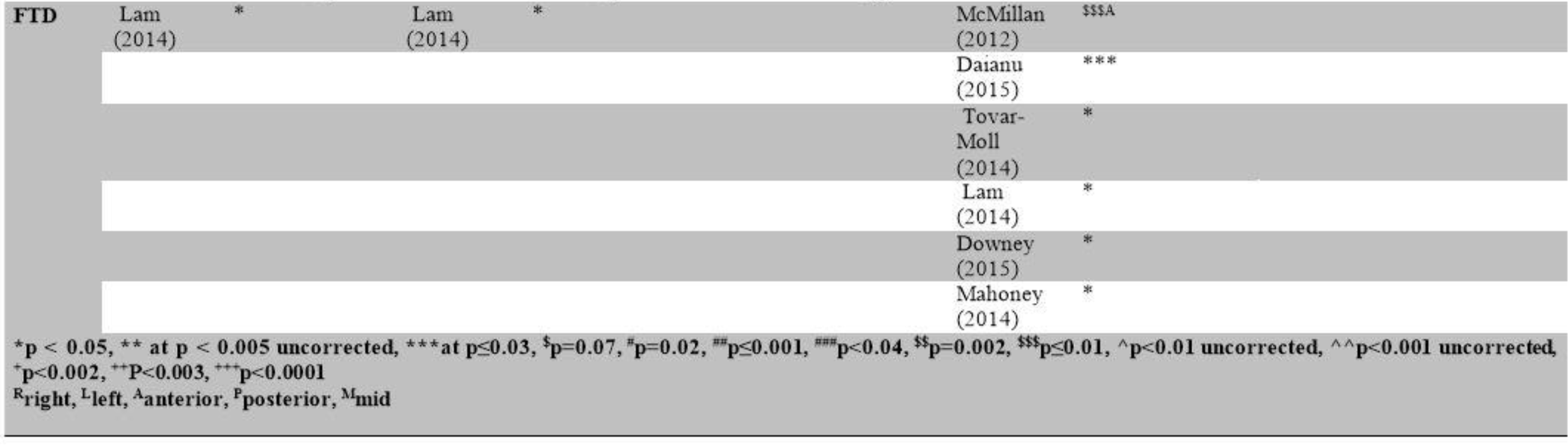
Studies of FA in the association fibersiwhich studied the association fibers.

There were widespread changes in the association fibers in ALS. Sarro et al found increased MD, AD and RD bilaterally in the SLF but no changes in FA (Sarro, Agosta et al. 2011). ROI analysis revealed changes in DTI metrics such as MD in the uncF and decreased FA in ILF (Pettit, Bastin et al. 2012) and right SLF with increased RD and MD (Menke, Körner et al. 2014). Many studies found that reduced FA was greatest in SLF (Agosta, Caso et al. 2013) and IFOF (Borroni B and et al. 2007). When cognitively impaired ALS (ALSci) were compared to controls and non-impaired counterparts, increased RD and MD were observed in the SLF, IFOF and uncF bilaterally while reduced FA were seen only when ALSci were compared to controls (Kasper, Schuster et al. 2014). Tractography in ALS showed increased AD in uncF compared to controls (Christidi, Zalonis et al. 2014). In later stages of ALS, more association fibers were involved in DTI changes than in the early stages (Trojsi, Caiazzo et al. 2015).While one study found reduced generalized FA^2^ in the left SLF (Trojsi, Corbo et al. 2013), another study also studied the arcuate fasciculus but found no significant changes in FA, MD or RD (Bastin, Pettit et al. 2013).

The correlations of DTI measures with quantitative clinical measure in ALS are summarized in Supp Tables 4.2–4.6. Using ROI, RD in the right SLF correlated with ALSFRS while both FA and RD correlated with disease progression (Menke, Körner et al. 2014). In regression analysis, reduced FA in bilateral IFOF and ILF was shown to negatively correlate with Trail-making test scores while the right IFOF and ILF associated with Stroop test scores (Sarro, Agosta et al. 2011). Negative correlation of DTI metrics with UMN scores and Frontal Systems Behavior Scale (FrSBe) was found in the SLF (Trojsi, Corbo et al. 2013). Crespi found an association between specific emotional recognition and FA, MD and mode of anisotropy ^3^ along the right ILF and IFOF but positive correlation with the cumulative scores of single negative emotions (Crespi, Cerami et al. 2014).

###### 5.1.1.2.3 Subcortical WM (Limbic tracts)

Studies showing significant and non-significant changes in DTI metrics in limbic tracts are presented in Table 4_6 and Figure 4.2.

**Table 4.6:**
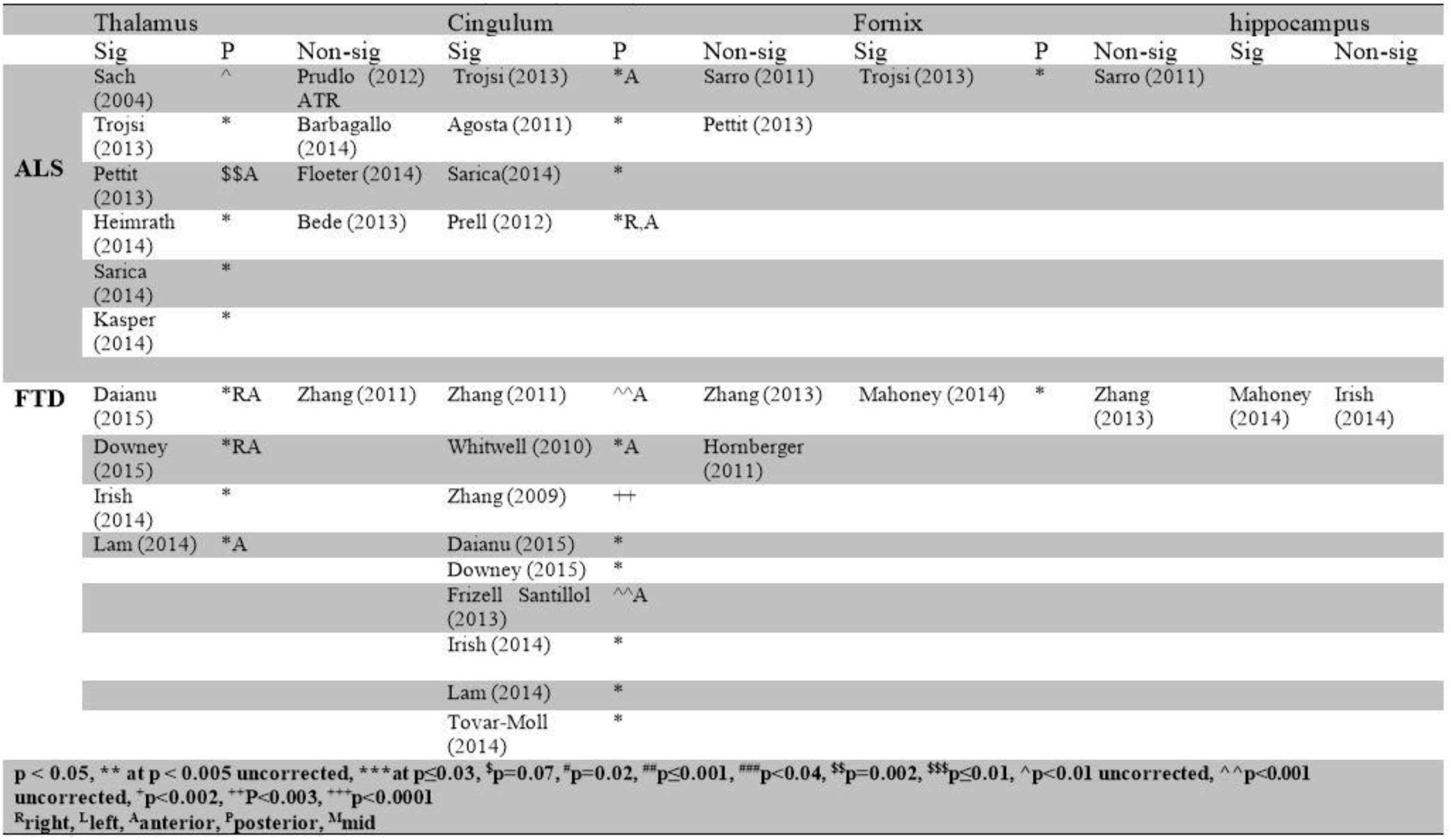
Stuclies of FA in the subcortical (limbic) tracts, which stucliecl the subcortical tracts.

There were widespread changes in the limbic tracts in ALS. There was reduced FA in the thalamus (Pettit, Bastin et al. 2012, Heimrath, Gorges et al. 2014), more significant on the right side (Sach, Winkler et al. 2004) and anteriorly (Trojsi, Corbo et al. 2013, Sarica, Cerasa et al. 2014). The hippocampus, thalamus and anterior cingulum were found to have increased RD (Cardenas-Blanco, Machts et al. 2014) and MD (Barbagallo, Nicoletti et al. 2014) in ALS specifically when using ROI-based methods (Kasper, Schuster et al. 2014), which was found to be correlated with reverse digital span (Pettit, Bastin et al. 2012). The average FA of the fornix was also reported to be reduced (Trojsi, Corbo et al. 2013). The amygdala was found by one group to have increased MD, which was negatively correlated with ALSFRS-R (Barbagallo, Nicoletti et al. 2014). ROI analysis showed significant changes in FA and MD in the thalamic radiation in the frontal region (Pettit, Bastin et al. 2012). Agosta et al reported similar changes when comparing ALS versus primary lateral sclerosis (PLS) (Agosta, Caso et al. 2013). One study found widespread reduction in FA and increase in MD in WM subcortical structures (Floeter, Katipally et al. 2014). In ALS patients at stage 3, where a third body region is involved, DTI metrics showed significant changes in thalamic radiation (ThR) (Trojsi, Caiazzo et al. 2015). When comparing ALS with their counterparts who had cognitive impairment, increased RD and MD were seen in ThR and Cg in ALSci (Kasper, Schuster et al. 2014).

The results of clinical correlation of DTI measures with clinical features of ALS in subcortical white matter tracts are summarized in Supp Tables 4.2–4.6. Modified Card Sorting Test (MSCT) and frontal assessment battery (FAB) scores were found to correlate with MD in WM tracts in caudate nucleus, amygdala and hippocampus in ALS (Barbagallo, Nicoletti et al. 2014). FA and MD in the fornix correlated with verbal learning and memory test scores (Filippi, Agosta et al. 2011). In the parahippocampal tract and the left thalamic radiation, increased FA correlated with increased apathy score on Frontal Systems Behavior Scale (FrSBe) (Tsujimoto, Senda et al. 2011). MD in the thalamus correlated positively with disease duration and negatively with ALSFRS-R (Barbagallo, Nicoletti et al. 2014).

###### 5.1.1.2.4 Commissural fibers

Studies showing both significant and non-significant changes in DTI metrics in commissural fiber tracts are presented in Table 4_7 and Figure 4.2.

**Table 4.7:**
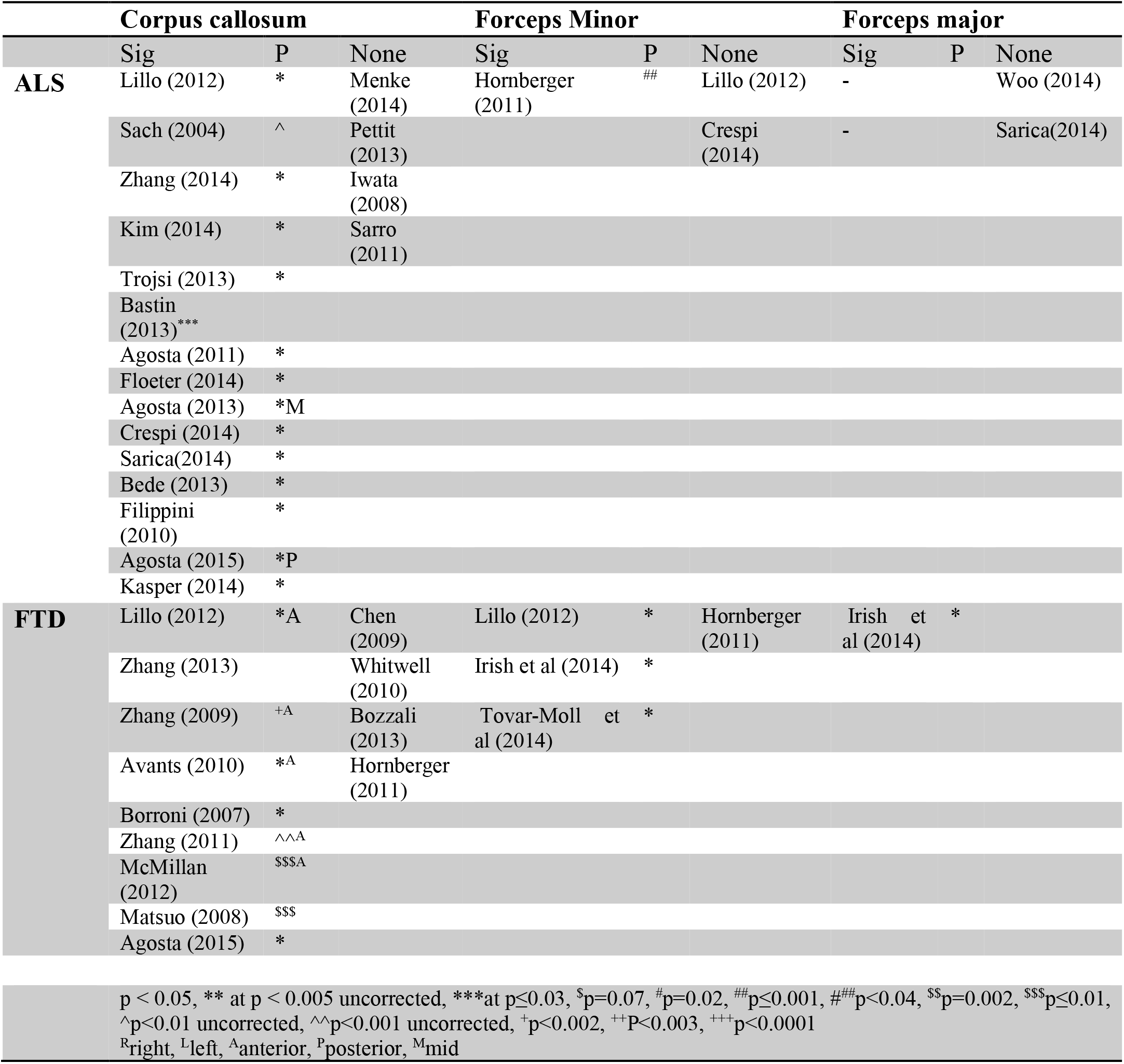
Studies of FA in the callosal radiation, which studied the callosal tracts.

Widespread changes in commissural fibers were reported. Reduced FA in the CC was found when comparing ALS and ALS-FTD group with controls (Lillo, Mioshi et al. 2012). Others reported the reduction in FA to be more dominant in the genu (Bastin, Pettit et al. 2013, Bede, Bokde et al. 2013) of the CC, forceps minor and major (Prudlo, Bissbort et al. 2012) but the body of the CC showed more changes when classic ALS patients were compared with controls (Prudlo, Bissbort et al. 2012, Agosta, Galantucci et al. 2015, Tang, Chen et al. 2015). Most studies found reduced FA in the CC (Sach, Winkler et al. 2004, Kasper, Schuster et al. 2014).

Increased MD (Sarro, Agosta et al. 2011, Trojsi, Caiazzo et al. 2015, Vora, Kumar et al. 2016) and RD (Kasper, Schuster et al. 2014, Zimmerman-Moreno, Ben Bashat et al. 2015) were reported in few studies. Prudle et al found that both forceps major and minor showed reduced FA in all ALS subtypes when using ROI analysis (Prudlo, Bissbort et al. 2012). When ROI is used, the genu and splenium of CC reflected significant reduction in the FA in ALS group (Vora, Kumar et al. 2016).

When not corrected for multiple comparisons, reduced FA showed weaker correlation with ALSFRS-R, but higher RD correlated with clinical scores such as UMN scores and ALSFRS-R (Filippini, Douaud et al. 2010). No correlations was found in the CC with ALSFRS-R or UMN scores (Filippini, Douaud et al. 2010). Sarro et al found that the changes in FA and MD in the CC correlated with the results of neuropsychological tests such as the Trail-making test scores and Stroop test which also correlated with the FA in the CC (Sarro, Agosta et al. 2011).

###### 5.1.1.2.5 Brain stem tracts

Studies showing significant and non-significant results in DTI metrics in brain stem tracts are presented in Table 4_8 and Figure 4.2.

**Table 4.8:**
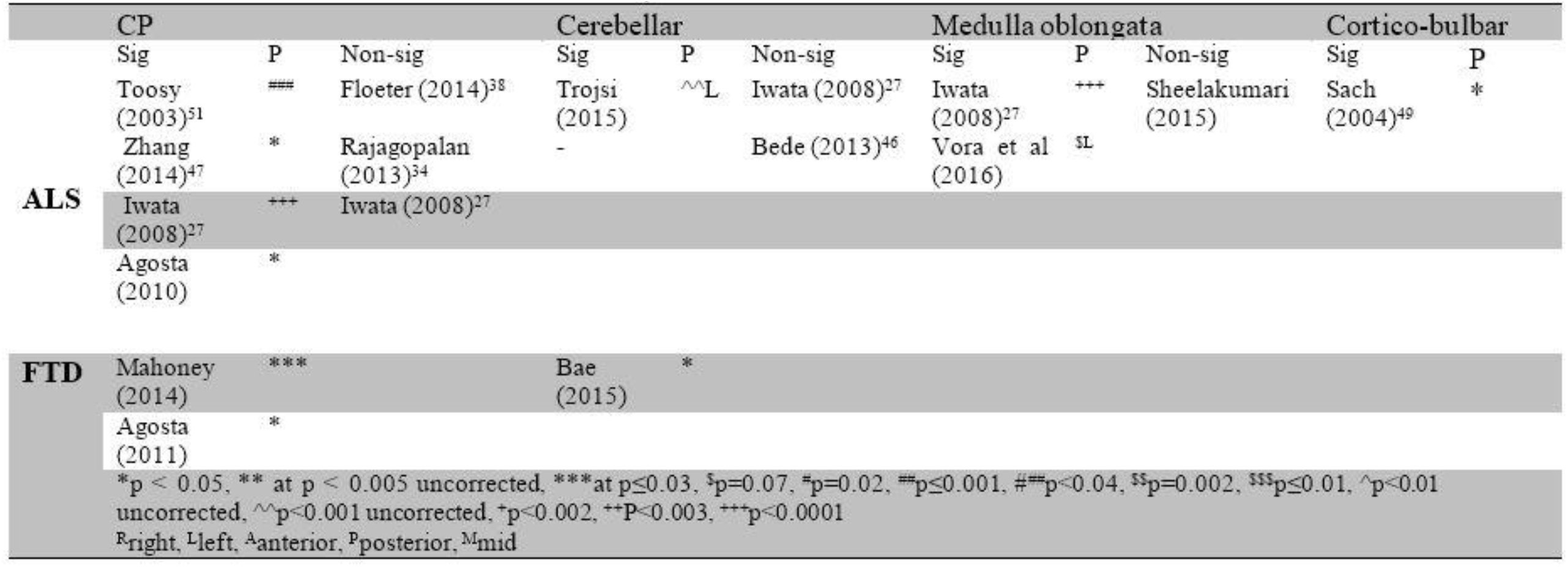
Stuclies of FA in the brain stem tracts which stucliecl the brain stem tracts.

Significant changes in FA were found in most of brainstem tracts; medulla oblongata (MD but not FA) (Sheelakumari, Madhusoodanan et al. 2015), cerebral peduncle (Sheelakumari, Madhusoodanan et al. 2015, Tang, Chen et al. 2015), and basis pontis (Iwata, Aoki et al. 2008). Reduced FA was reported in the left medullary pyramid (Vora, Kumar et al. 2016) and bilaterally in cerebellar peduncle (CP) (Tang, Chen et al. 2015) in ALS compared to controls. Bede et al divided ALS subjects based on results of testing for C9orf72 hexanucleotide repeats (C9+ve and C9–ve) and found that WM exhibited changes in the cerebellar pathways in both groups (Bede, Bokde et al. 2013). In another study comparing ALS versus primary lateral sclerosis (PLS), increased MD was found in the bilateral middle CP (Sheelakumari, Madhusoodanan et al. 2015) and transverse pontine fibers (Floeter, Katipally et al. 2014). Reduced FA in the medulla oblongata correlated with higher UMN scores in ALS patients (Iwata, Aoki et al. 2008).

#### 5.1.2 DTI in FTD

A summary of the number of papers that studied each tract, and the number of these papers that found significant abnormalities in each tract using FA is provided in Table 4_3. A visual summary of the WM tracts that found to be significant in FTD studies is presented in Figure 4.3.

##### 5.1.2.1 Whole brain analysis

Nine whole-brain studies of FTD were included (Zhang, Schuff et al. 2009, Agosta, Scola et al. 2011, Hornberger, Geng et al. 2011, Lillo, Mioshi et al. 2012, Zhang, Tartaglia et al. 2013, Lam, Halliday et al. 2014, Tovar-Moll, de Oliveira-Souza et al. 2014, Agosta, Galantucci et al. 2015, Downey, Mahoney et al. 2015). When FTD patients were compared to ALS and ALS-FTD patients, TBSS revealed significant changes involving the callosal tracts; the forceps minor, and ILF than in CST (Lillo, Mioshi et al. 2012). Reduced FA and increased MD were also found in CC, orbitofrontal, occipital and frontoparietal WM in behavioral variant FTD (bvFTD) subjects when compared to controls (Agosta, Scola et al. 2011, Agosta, Galantucci et al. 2015). The most pronounced white matter degeneration in FTD patients was seen in the anterior callosal region (Zhang, Schuff et al. 2009). Increases in RD and AD were similar and shown to be sharing the same regions when FTD were compared to controls. These regions included; anterior callosal region, fornix (Agosta, Scola et al. 2011) and bilateral uncF (Zhang, Schuff et al. 2009). Changes in FA and AD in uncF were found when comparing FTD versus healthy controls (Zhang, Schuff et al. 2009). It has been reported that FA changes were mostly dorsal and ventral when comparing bvFTD versus controls in voxel-wise analysis (Tovar-Moll, de Oliveira-Souza et al. 2014) and whole brain TBSS (Downey, Mahoney et al. 2015). Longitudinal study have documented that DTI changes involved the callosal, association and cingulate tracts in bvFTD and that WM changes overlap GM atrophy (Lam, Halliday et al. 2014). Reduced FA correlated negatively with Hayling task and neuropsychiatric inventory disinhibition in UncF (Hornberger, Geng et al. 2011). Abnormalities in CR and ALIC were also reported when using voxel-wise analysis (Tovar-Moll, de Oliveira-Souza et al. 2014). External capsule and IC were found to have reduced FA but increased RD (Agosta, Scola et al. 2011). Some studies reported that RD shows more regions than other DTI metrics in FTD group (Zhang, Schuff et al. 2009, Lam, Halliday et al. 2014).

##### 5.1.2.2 ROI and Tractography analysis

###### 5.1.2.2.1 Projection Fibers

Table 4_4 shows all the studies that involved projection fibers. Decreased FA bilaterally in the overall CST was reported in two studies (Lillo, Mioshi et al. 2012, McMillan, Brun et al. 2012), but more specifically in the left in one study (Borroni B and et al. 2007). FA was found to be reduced at the level of both cerebral peduncles (CP) (Agosta, Scola et al. 2011, Mahoney, Beck et al. 2012) and corona radiata (CR) with increased AD (Borroni B and et al. 2007, Agosta, Scola et al. 2011) and RD on the right CST (Zhang, Schuff et al. 2009). However, other studies reported increased FA along the whole CST (Avants, Cook et al. 2010, Whitwell, Avula et al. 2010).

Although severe degeneration across the motor system was found when comparing bvFTD with controls and ALS patients (Bae, Ferguson et al. 2016), another study reported the CST to be the least affected tract in bvFTD (Daianu, Mendez et al. 2015).The only study that addressed DTI correlations showed that in FTD, reduced FA correlated with cortical thickness (Avants, Cook et al. 2010).

###### 5.1.2.2.2 Association Fibers

Table 4_5 shows FTD studies that included association fibers. Reduced FA was reported in all papers that investigated the SLF (Borroni B and et al. 2007, Avants, Cook et al. 2010, Whitwell, Avula et al. 2010, McMillan, Brun et al. 2012). This reduction in FA was associated with increased RD in both the SLF and ILF particularly in the anterior regions (Whitwell, Avula et al. 2010). Reduced FA was also reported in the right SLF in fontal variant FTD (fvFTD) and left SLF in temporal variant FTD (tvFTD), bilaterally in ILF and IFOF when compared to controls while changes were bilateral in the ILF and IFOF (Borroni, Brambati et al. 2007). Changes in uncF were mostly frontal and temporal when comparing bvFTD versus controls in ROI (Tovar-Moll, de Oliveira-Souza et al. 2014).

When comparing bvFTD versus controls, DTI revealed abnormalities in most of the association fibers bilaterally (Irish, Hornberger et al. 2014, Tovar-Moll, de Oliveira-Souza et al. 2014, Daianu, Mendez et al. 2015). This was noted even with longitudinal studies that showed changes of DTI metrics at base line in regions of ILF, IFOF and uncF (Mahoney, Simpson et al. 2015). SLF and ILF had increased MD and AD after 12 months (Lam, Halliday et al. 2014). Rate of changes^4^ in DTI metrics in uncF were significant when bvFTD compared to controls in a longitudinal study (Mahoney, Simpson et al. 2015).

WM changes in the SLF, IFOF, uncF and ILF correlated with cortical thickness in FTD (Avants, Cook et al. 2010). A negative correlation was reported between FA in SLF and behavioural and neuropsychological examinations (Borroni, Brambati et al. 2007). Remote ABM retrieval correlated significantly with FA in the left uncF (Irish, Hornberger et al. 2014). DTI measures in the uncF correlated with NPI-aberrant motor behavior sub-score especially in the frontal part (MD and FA) and temporal part (FA) (Tovar-Moll, de Oliveira-Souza et al. 2014), Supp Table 6.

###### 5.1.2.2.3 Subcortical WM (Limbic tracts)

All FTD studies that included limbic tracts are shown in Table 4_6. Reduced FA and increased MD were found in the frontotemporal subcortical WM structures such as fornix, anterior ThR (Irish, Hornberger et al. 2014) and cingulum (Floeter, Katipally et al. 2014, Irish, Hornberger et al. 2014, Tovar-Moll, de Oliveira-Souza et al. 2014), but significant changes in the latter were seen only in FA in the bvFTD group when compared to controls (Agosta, Scola et al. 2011).

When using TOI, the fornix exhibited changes in FA, AD and MD (Zhang, Tartaglia et al. 2013). One study reported increased RD and AD in parahippocampal cingulum (Zhang, Tartaglia et al. 2013). Right parahippocampal cingulum showed changes in all assessed DTI metrics in a cross-sectional DTI study that compared bvFTD and controls (Mahoney, Simpson et al. 2015). MD was reported to be increased in ThR and Cg in bvFTD group compared to controls (Daianu, Mendez et al. 2015). At baseline, when comparing bvFTD and controls, all DTI measures reflected significant changes in the anterior ThR and Cg but this was not reported after 12 months (Lam, Halliday et al. 2014). FA changes in the right anterior ThR and fornix had positive correlation with cognitive performance in bvFTD patients (Downey, Mahoney et al. 2015). In bvFTD, DTI changes in the posterior Cg, the parahippocampal part (paraHpc), correlated with NPI-apathy subscores (MD, p<0.01), and Mattis total scores (MD and FA, p<0.05) (Tovar-Moll, de Oliveira-Souza et al. 2014).

###### 5.1.2.2.4 Commissural fibers

FTD studies that investigated commissural fibers are shown in Table 7. There was reduced FA in the CC (Matsuo, Mizuno et al. 2008, Avants, Cook et al. 2010, Whitwell, Avula et al. 2010, Lillo, Mioshi et al. 2012, McMillan, Brun et al. 2012) and more anteriorly when not corrected for multiple comparisons (Zhang, Schuff et al. 2011). Increased MD was reported when comparing a small group of seven FTD subjects with controls (Chen, Lin et al. 2009). Changes in DTI metrics were found in the CC in most of the fronto-temporal lobar degeneration (FTLD) subgroups (Agosta, Scola et al. 2011). However, in the bvFTD group, when compared to controls, there were changes in FA, MD and AD (Agosta, Scola et al. 2011). Reduced FA in left CC was reported in tvFTD when compared with healthy controls (Borroni, Brambati et al. 2007). At baseline, all DTI metrics exhibited significant changes in the genu of CC, while after 12 months the significant changes in FA and RD extended to bilateral splenium of CC (sCC) (Lam, Halliday et al. 2014). Similar results were reported in bvFTD with all DTI measures in the frontal WM including gCC while this was not observed in sCC (Lu, Lee et al. 2014). Others reported the DTI changes to be in the bCC, bilaterally, in both cross-sectional and longitudinal DTI studies (Lam, Halliday et al. 2014) but the rates of change in FA and MD at later stage involved the bCC for FA, RD, MD and AD only in the c9orf72 group. All assessed DTI metrics showed significant changes in the sCC (Mahoney, Simpson et al. 2015). Using tractography in bvFTD subjects, the genu of CC shows altered diffusion anisotropy (Daianu, Mendez et al. 2015). DTI changes in CC were mostly dorsal and ventral in bvFTD compared to controls (Downey, Mahoney et al. 2015). With ROI, FA and MD changes in sCC were found to be significant in FTD group compared to controls. Similar results were seen when using voxel-wise analysis (Tovar-Moll, de Oliveira-Souza et al. 2014).

Behavioral variables were found to be associated with DTI measures in gCC (Lu, Lee et al. 2014). Lillo et al found that FA changes in the anterior CC and forceps minor occurred in the bvFTD and ALS-FTD subjects when compared to other patient subgroups and controls but minimal changes in these regions were evident when comparing the pure ALS patients and healthy controls (Lillo, Mioshi et al. 2012). Within bvFTD group, impairment of emotion identification impairment was associated with WM alteration in CC (Downey, Mahoney et al. 2015).

###### 5.1.2.2.5 Brain stem tracts

A total of three FTD studies examined DTI changes in brain stem, Table 4_8. Significant changes in FA was found in the bvFTD group in the CP when compared with healthy controls only with FA but not with other DTI metrics (Agosta, Scola et al. 2011). The FTD group, who was C9ORF72 positive, had reduced FA in the superior CP compared to controls (Mahoney, Simpson et al. 2015).

#### 5.1.3 Overlap between ALS and FTD

This review aimed to determine the overlap between ALS and FTD, and also to demonstrates the differences between ALS and FTD. The SLF, ILF, IFOF, UncF, ThR, Cg, Fx and CC were found to be abnormal in 50% or more of the studies in ALS and FTD. There was strong evidence of involvement of uncF in FTD, but only a minority of studies found abnormality of uncF in ALS. There were abnormalities in the cingulum and IFOF in both ALS and FTD, and forceps minor was reported to be abnormal in minority of studies of ALS but more evident in FTD studies. As shown in Table 4, there is extensive evidence that the CST was abnormal in ALS, and less evidence in FTD, where there were fewer studies that only examined the average FA of the whole CST. However, two studies have reported CR to show significant changes in FTD.

Figures 4.2–4.4 show the tracts that were abnormal in ALS, FTD and in both ALS and FTD, respectively, using tracts that were abnormal in more than 66% of studies, where the evidence is strongest.

**Figure 4.2:**
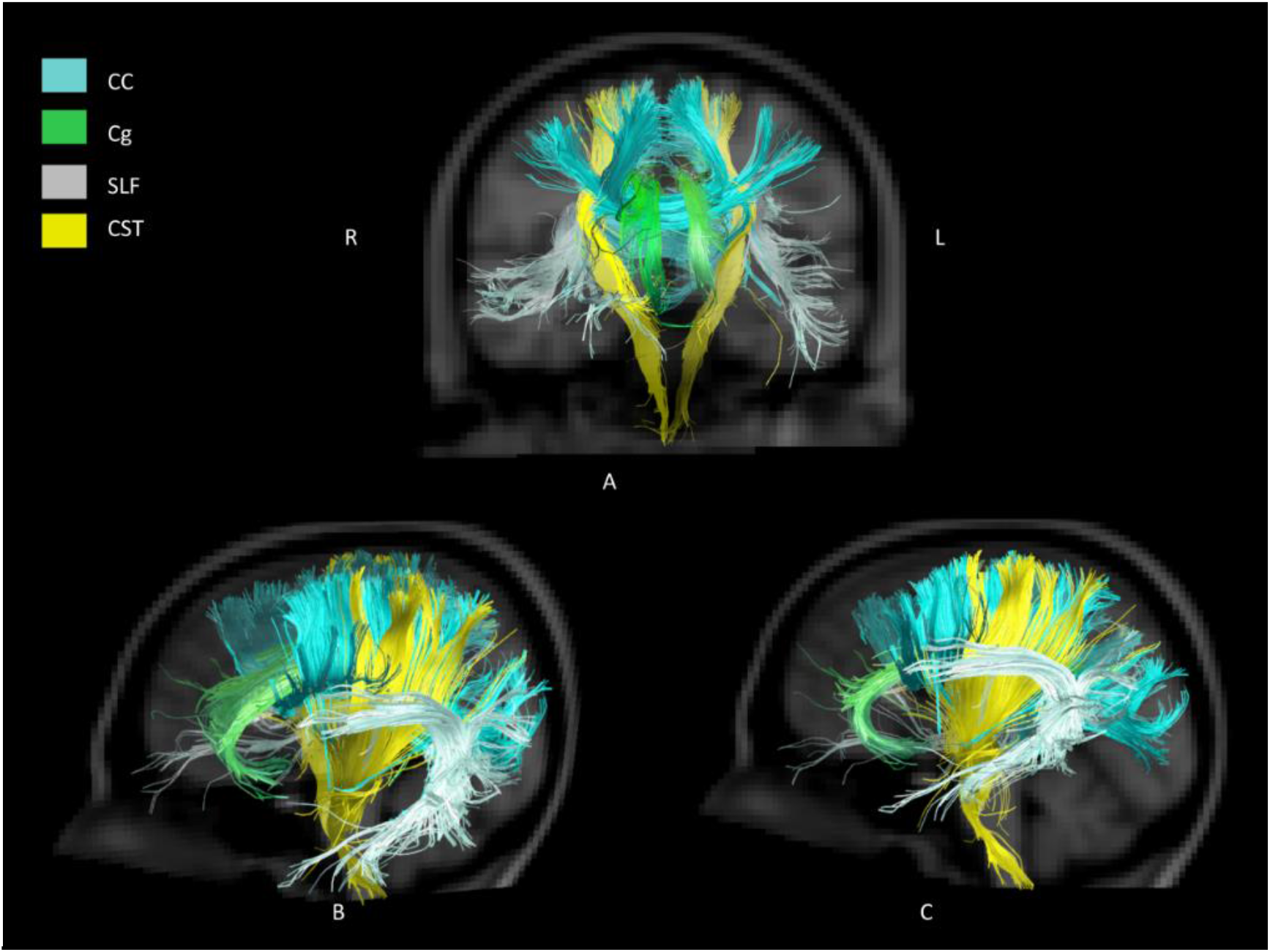
Visual representation of WM tracts that were found significant in ALS studies.

**Figure 4.3:**
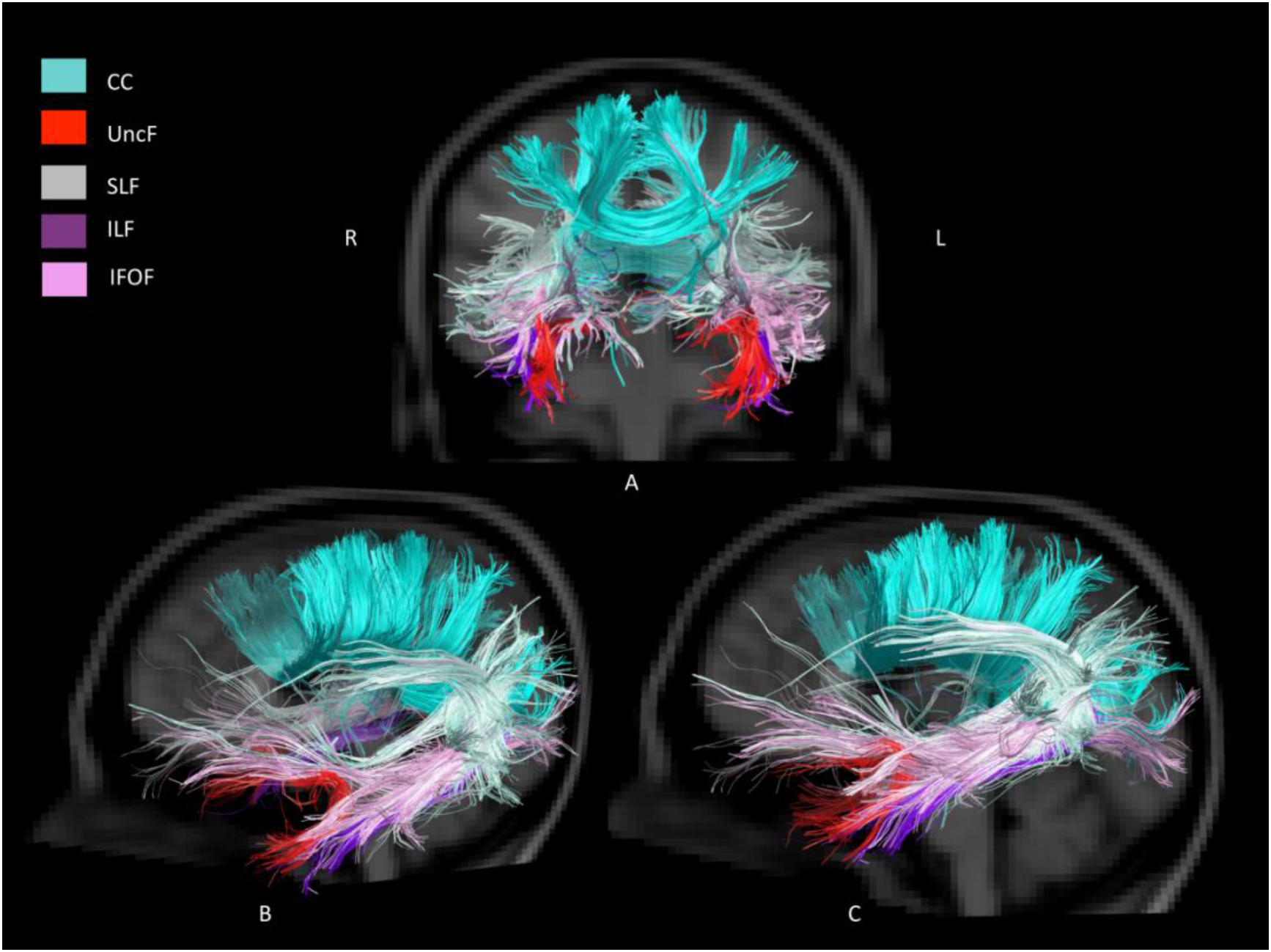
Visual representation of WM tracts that were found significant in FTD studies. (CST was removed for better visualization of other tracts).

**Figure 4.4:**
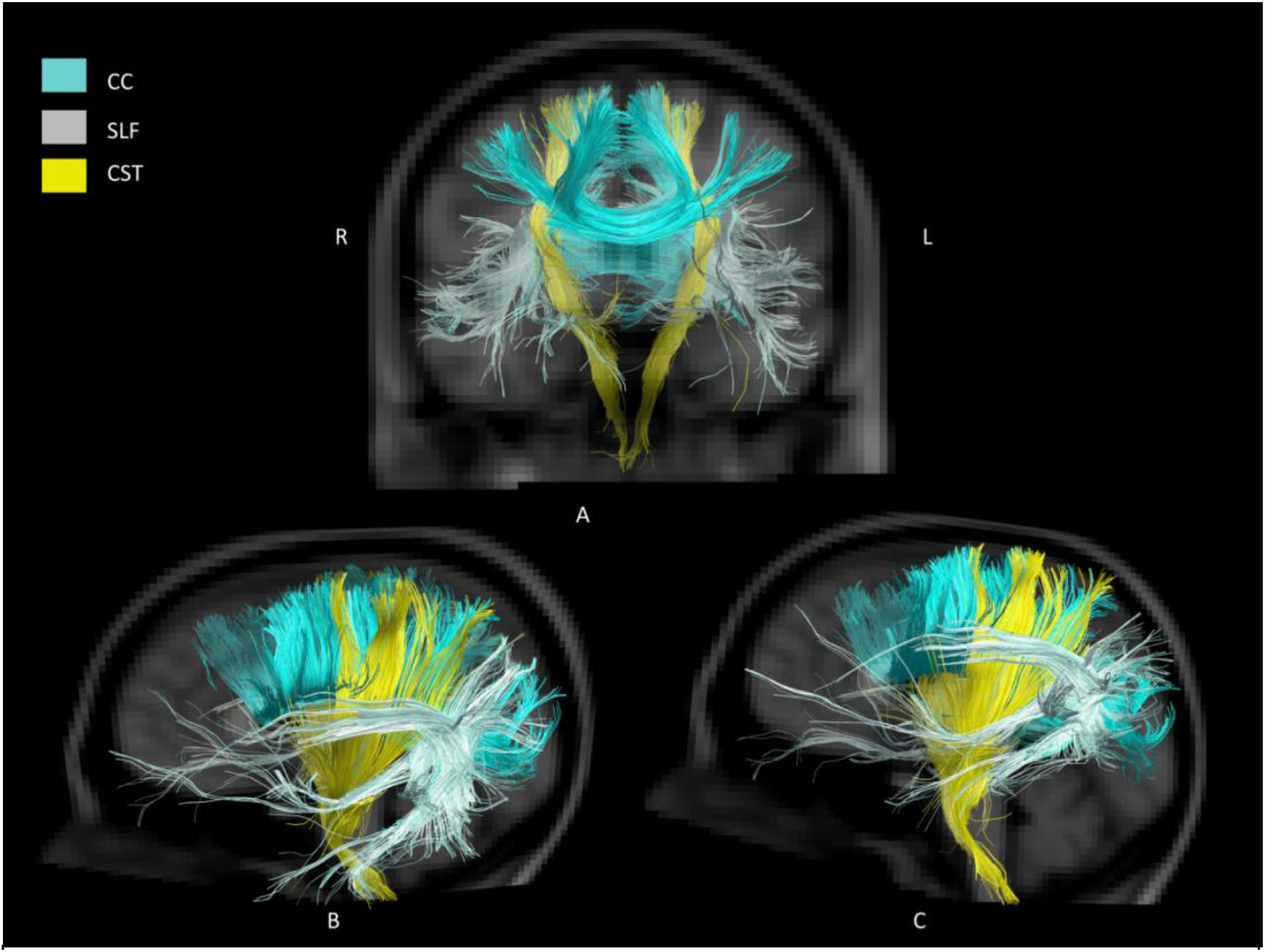
Visual representation of WM tracts that are found to be significant and shared in both ALS and FTD studies.

## 6 Discussion

DTI is a promising method to assess upper motor neuron dysfunction (Wang, Melhem et al. 2011, Turner, Agosta et al. 2012) and changes in frontal lobe function (Naik, Lundervold et al. 2010). We have systematically reviewed the abnormalities that are reported in ALS and in FTD using DTI. We report data obtained from 1674 patients across 64 studies. All studies have reported the results of FA; however, the other DTI measures were not consistently analyzed across all studies. Because there is an overlap of clinical features between ALS and FTD, we expected that there would be overlap of the white matter tracts involved. There have been previous meta-analyses of DTI abnormality in ALS (Foerster, Dwamena et al. 2012, Li, Pan et al. 2012). There are no previous systematic reviews of DTI in FTD. We were not able to obtain the raw data for most of these studies, and the studies varied in the use of sequences and in the techniques for analysis of the data, so we have made a comparison of the abnormalities that were reported.

DTI measures were variously obtained using ROI, TBSS, tractography and other methods. ROI is susceptible to inclusion of cerebrospinal fluid (CSF) or gray matter (GM) (Snook, Plewes et al. 2007). TBSS is considered to be the standard approach for voxel-based analysis (VBA) of DTI data (Smith, Jenkinson et al. 2006), but has drawbacks as described previously such as misalignment and alteration in the thickness of the skeleton (Edden and Jones 2011, Zalesky 2011, Keihaninejad, Ryan et al. 2012, Bach, Laun et al. 2014). Tractography is unable to determine the precise origin and termination of tracts, but can guide the ROI analysis of WM (Gong, Jiang et al. 2005, Yushkevich, Zhang et al. 2008, Goodlett, Fletcher et al. 2009, Jbabdi, Behrens et al. 2010). Because of the limited number of studies, we have combined the results of all the studies despite their different technical approaches, as outlined in the extracted data table in the appendix. However, we noted that some older studies were limited in resolution, power and field strength. This could account for some of the discrepancies. The tracts that showed significant changes are displayed in Figures 4.2 to 4.4.

### 6.1 ALS studies

The most common finding in ALS is the reduction in FA in the CST. Classical pathological studies of ALS show loss of upper and lower motor neurons and sclerosis of the CST in the spinal cord (Rafałowska and Dziewulska 1995). Other post mortem studies found gliosis affecting the posterior third of the internal capsule (Abe, Fujimura et al. 1997). Animal studies found similar results in cerebral motor cortex, where the CST originates (Martin 1999). Thus, abnormality of the CST in DTI of ALS studies is an expected finding. DTI studies also showed extensive abnormality of the CC, the cingulum and thalamus, confirming the findings of a previous review (Li, Pan et al. 2012).

The abnormalities found in ALS with DTI are consistent with the results of the imaging studies using other techniques. For example, structural MRI studies in ALS indicated that hyperintensities in CST and hypointensity in motor cortex are consistent features (Hecht, Fellner et al. 2001, Zhang, Uluǧ et al. 2003). Analysis of regional atrophy in VBM studies in ALS patients suggests that the GM loss occurs in the motor cortex and frontal, temporal and limbic areas (Chang, Lomen-Hoerth et al. 2005, Kassubek, Unrath et al. 2005, Mezzapesa, Ceccarelli et al. 2007). Susceptibility weighted imaging (SWI) has also found abnormalities of these pathways in ALS (Prell, Hartung et al. 2015). Functional MRI (fMRI) in ALS patients has revealed that impaired letter fluency correlated with impaired activation in the frontal and cingulate gyri (Abrahams, Goldstein et al. 2004). Positron emission tomography (PET) also found reduced regional perfusion in the sensorimotor cortex, and also extra-motor areas such as prefrontal, cingulate, putamen and parietal cortices (Hatazawa, Brooks et al. 1988, Habert, Lacomblez et al. 2007) and that reduction is accompanied by mild neuropsychological deficit (Ludolph, Langen et al. 1992).

In ALS, many of the DTI studies show positive correlation between FA and performance on some cognitive examinations such as Trail-making, Stroop test scores and letter fluency. These changes are mainly in the projection fibers, commissural tract and association fibers. However, negative correlation was found with emotional recognition in the association fibers. Verbal learning, memory test and the apathy scores on FsRBe correlated with FA in the limbic tracts.

Our results found that the clinical measures positively correlated with the disease duration and disease progression but negatively with the UMN scores. ALSFRS-R had weaker correlation with FA in the commissural and association fibers but stronger negative correlation with MD in the CST and thalamic radiation. This is in support of the theory that ALS involves more than just motor dysfunction. Previous meta-analysis in ALS using DTI measures is in support of the findings in this review (Li, Pan et al. 2012).

### 6.2 FTD studies

The tracts that were abnormal in FTD included the SLF, ILF and IFOF as well as the corpus callosum and uncinated fasciculus. There also was abnormality of the CST. FTD is associated with neurodegeneration and atrophy of the frontal, parietal and temporal lobes (Whitwell, Jack et al. 2007). In a previous post-mortem DTI study, histopathologic correlation revealed changes and frontal degeneration with mild frontotemporal gliosis (Larsson, Englund et al. 2004). FTD subtype studies reported the involvement of association fibers in the disease prognosis particularly ILF, IFOF and SLF, and the callosal radiation in the left hemisphere in tvFTD.

Other MRI studies of FTD patients have shown patterns of GM loss primarily in the frontal, orbitofrontal (Harlow 1868, Hornberger, Geng et al. 2011) and anterior temporal lobes (Baron, Chetelat et al. 2001, Diehl, Grimmer et al. 2004, Grimmer, Diehl et al. 2004, Ishii, Sasaki et al. 2005, Jeong, Cho et al. 2005). Functional studies have shown the changes to be prominent in the parietal lobe and the cingulate gyrus (Bartenstein, Minoshima et al. 1997, Johnson, Jahng et al. 2006). Although several researchers have discovered a contribution by the association tracts in disease progression of FTD, less is known about the tract-specific contribution in cognition and this has not been fully explored (Cairns, Bigio et al. 2007).

### 6.3 Overlap between ALS and FTD

The pathways for which there is strong evidence of abnormality in both ALS and FTD are shown in Figures 4.2–4.4. It is thought that ALS and FTD lie on a spectrum and that subjects with the combination of ALS and FTD have worse survival than subjects with ALS without FTD (Mathuranath, Nestor et al. 2000). This review identified extensive evidence of CST abnormalities in ALS and also some evidence in FTD. The DTI studies show extra-motor involvement in ALS and FTD in the cingulum and the CC, especially the forceps minor which reflects the neuropathological findings of degeneration in the CC (Brownell, Oppenheimer et al. 1970) and corresponds to the severe atrophy of the most anterior CC that has been related to cognitive impairment (Yamauchi, Fukuyama et al. 1995). Earlier studies showed that a network of orbitofrontal, anterior temporal and mesial frontal brain regions and their connecting white matter tracts are involved in ALS and FTD (Hornberger, Geng et al. 2011, Mahoney, Beck et al. 2012). The integrity of WM tracts connecting these regions, namely uncF, forceps minor and callosal radiation appears critical. The atrophy of frontal regions is important as it is involved in Theory of Mind processing (Stone, Baron-Cohen et al. 1998, Ferstl and von Cramon 2002) and cognitive control (Miller 2000, Ridderinkhof, Ullsperger et al. 2004). Involvement of the cingulate region in cognition is well established (Bush, Luu et al. 2000). Moreover, association fibers such as uncF (Agosta, Pagani et al. 2010) which is involved in naming and language function, (Papagno 2011) and SLF (Abrahams, Goldstein et al. 2005) were involved in ALS. Several of the studies have reported increased AD of the uncF in ALS and suggested that this may relate to the behavioral disturbances of ALS patients.

Association tracts are also involved in cognition. In Alzheimer disease there is loss of integrity of the association fibers (Rose, Chen et al. 2000), and WM destruction and altered regional morphology of CC (Black, Moffat et al. 2000, Meng, Guo et al. 2012). Efficient organization of association fibers is essential for optimal cognitive performance (Schmithorst, Wilke et al. 2002, Frye, Hasan et al. 2010). There is evidence that emotional deficit follows selective damage in the ILF and IFOF (Philippi, Mehta et al. 2009). The role of the corticostriatal pathways in neurodegeneration has recently been emphasized (Shepherd 2013), so the involvement of subcortical area may underlie the cognitive and behavioral changes in ALS (Tsermentseli, Leigh et al. 2012).

The heterogeneity of impairment in ALS is well-studied and includes cognitive deficits encompassing language and memory impairment specifically the verbal fluency (Raaphorst, De Visser et al. 2010, Mackenzie, Ansorge et al. 2011). One complicating factor in most of the included studies is that some ALS subjects did not undergo cognitive testing. It is possible that the regions of abnormality that we have found to overlap between ALS and FTD are due to these abnormalities being present only in ALS subjects with cognitive impairment.

In summary, this systematic review provides valuable insights into the DTI abnormalities in ALS and FTD, emphasizing commonalities and differences between these conditions, particularly in white matter tracts and their implications for cognitive functions. It underscores the potential of DTI as a tool for understanding the pathophysiology of these diseases and highlights the need for standardized DTI analysis in future clinical studies. DTI metrics demonstrate the significant differences between controls and patients and may be used to predict disease progression. Further investigations are needed to identify the overlap of ALS and FTD in cognitive processes to address the progression of the disease. Although DTI findings are important to the scientific findings, to date, they are not readily transferrable to clinical practice. Standardization of DTI analysis is necessary to allow for clinical studies, which is still a challenge due to the rapid advances in DTI techniques.

## Supporting information

Supplementary table of extracted data

## Data Availability

All data produced in the present study are available upon reasonable request to the authors

## Footnotes

1 Index of structural connectivity (CI) is the proportion of plausible paths connecting two ROIs, averaged across tractography directions.

6 The generalized fractional anisotropy (GFA) was calculated as GFA = std(ψ)/rms(ψ), where std is the standard deviation and rms is the root-mean-square.

5 Mode of anisotropy is a recently developed measure of anisotropy providing information about the shape of the tensor.

6 Was calculated based on the time from baseline scan in years, and interaction between disease group and time included to provide estimates of differences in the rate of change as a percentage per year.

