## Supplementary table of extracted data for "Unraveling Shared Pathways: A Comprehensive Systematic Review of Common Fiber Tracts in Amyotrophic Lateral Sclerosis and Frontotemporal Dementia using Diffusion Tensor Imaging"

### Appendix

INCLUSION CRITERIA:

- ≥ 18 y.o.
- El Escorial &/or Neary criteria
- Adequate descriptions of sample (M/F, age, other neuro dx)
- DTI tractography or ROI for WM tracts (ALS or FTD vs. HC)
- Adequate description of imaging protocol (MR sequence, b-value, diffusion directions, threshold) & stats (mean, SD)
- Measured FA and MD values in specific WM tracts with defined coordinates

QUADAS CRITERIA:

| Item | | Yes | No | Unclear |
|---|---|---|---|---|
| 1. | Was the spectrum of patients representative of the patients who will receive the test in practice? | ( ) | ( ) | ( ) |
| 2. | Were selection criteria clearly described? | ( ) | ( ) | ( ) |
| 3. | Is the reference standard likely to correctly classify the target condition? | ( ) | ( ) | ( ) |
| 4. | Is the time period between reference standard and index test short enough to be reasonably sure that the target condition did not change between the two tests? | ( ) | ( ) | ( ) |
| 5. | Did the whole sample or a random selection of the sample, receive verification using a reference standard of diagnosis? | ( ) | ( ) | ( ) |
| 6. | Did patients receive the same reference standard regardless of the index test result? | ( ) | ( ) | ( ) |
| 7. | Was the reference standard independent of the index test (i.e. the index test did not form part of the reference standard)? | ( ) | ( ) | ( ) |
| 8. | Was the execution of the index test described in sufficient detail to permit replication of the test? | ( ) | ( ) | ( ) |
| 9. | Was the execution of the reference standard described in sufficient detail to permit its replication? | ( ) | ( ) | ( ) |
| 10. | Were the index test results interpreted without knowledge of the results of the reference standard? | ( ) | ( ) | ( ) |
| 11. | Were the reference standard results interpreted without knowledge of the results of the index test? | ( ) | ( ) | ( ) |
| 12. | Were the same clinical data available when test results were interpreted as would be available when the test is used in practice? | ( ) | ( ) | ( ) |
| 13. | Were uninterpretable/ intermediate test results reported? | ( ) | ( ) | ( ) |
| 14. | Were withdrawals from the study explained? | ( ) | ( ) | ( ) |

| Study | Subjects | T | DTI Measures | Sequences & Analysis | Smoothing | Directions | B value | FA Thresh | WM Regions | Extracted Data | QUADAS |
|---|---|---|---|---|---|---|---|---|---|---|---|
| Toosy et al (2003) | 21 ALS<br>14 HC | 1.5 | Averages<br>FA<br>MD | Single-shot SE EPI<br><br>DispImage (manual ROI)<br><br>SAS 6.12 PROC MIXED used for stats | Not stated | 7 non-collinear directions | Max 700 s/mm2 | Not stated | CST at 4 levels (internal capsule, peduncles, pons, pyramids) | • Results all presented in graphs<br>• ↓ trend in FA<br>○ From peduncles to pyramids (p = 0.0001), but no different in ALS and HC (p = 0.48)<br>○ Mean FA is lower in ALS compared to HC at IC (p=0.036), CP, pons and pyramids (p=0.038)<br>• ↓ mean FA in ALS than HC at all levels (results given for midpoint 55 y.o.):<br>○ Lt. IC mean difference in FA = 0.049 (p = 0.032); rt. IC mean difference in FA = 0.058 (p = 0.011)<br>○ Lt below IC mean difference in FA = 0.052 (p = 0.001); rt. below IC mean difference in FA = 0.042 (p = 0.01)<br>• ↑ trend in MD from IC to pyramids<br>○ Significant difference in MD between ALS and HC at Left IC only (p = 0.01) | 1. Yes<br>2. No (no exclusion criteria)<br>3. Yes<br>4. Yes (duration included in formula)<br>5. Yes<br>6. Yes<br>7. Yes<br>8. Yes<br>9. Yes<br>10. No (El Escorial already known)<br>11. Yes<br>12. Yes<br>13. Unclear<br>**14.** Unclear (no withdrawals) |
| Abe et al (2004) | 7 ALS<br>11 HC | 1.5 | FA<br>MD | Single-shot SE EPI<br><br>dTV software (2-ROI methods)<br><br>SPM99; Matlab 5.3; | 8mm FWHM, then 12mm FWHM Gaussian kernel | 6 non-collinear directions | 1000 s/mm2 | FA = 0.1 | CST | • ↓ FA in ALS compared to HC<br>○ Rt. frontal subgyral WM ↓ FA (p < 0.007)<br>○ Lt. frontal preCG (p < 0.042)<br>• Tractography corresponded to ↓ FA in average CST<br>• No exact values for FA provided (visual only) | 1. Unclear (small numbers)<br>2. Yes<br>3. Yes<br>4. Unclear<br>5. Yes<br>6. Yes<br>7. Yes<br>8. Yes<br>9. Yes<br>10. No (El Escorial already known)<br>11. Yes<br>12. Yes<br>13. Unclear<br>14. Unclear (no withdrawals) |
| Karlsborg et al (2004) | 8 ALS<br>11 HC | 1.5 | FA<br>ADC | SE-DWI-EPI sequence<br><br>ROI (on T2-WI then aligned to DTI) | Not stated | 6 directions | 550 s/mm2 | Not stated | CST in 3 regions (corona radiate (CR), internal | • Overall ADC of CST higher than in HC (p = 0.04) and overall FA lower than in HC (p = 0.01); (no raw values given)<br>○ CR ADC = 0.80 ± 0.03 (p = 0.66 vs HC)<br>○ IC ADC = 0.79 ± 0.02 (p = 0.02 vs HC) | 1. Unclear (small numbers)<br>2. Yes<br>3. Yes<br>4. Yes (El Escorial applied at time of MRI)<br>5. Yes<br>6. Yes |

| | | | | | | | | | | | |
|---|---|---|---|---|---|---|---|---|---|---|---|
| | | | | MRI software not stated; BMDP and Minitab used for stats | | | | | capsule (IC), pons) | ○ Pons ADC = 0.79 ± 0.04 (p = 0.06 vs HC)<br>○ CR FA = 0.44 ± 0.06 (p = 0.48 vs HC)<br>○ IC FA = 0.59 ± 0.04 (p = 0.06 vs HC)<br>○ Pons FA = 0.57 ± 0.03 (p = 0.03 vs HC) | 7. Yes<br>8. Unclear (software not stated)<br>9. Yes<br>10. No (El Escorial already known)<br>11. Yes<br>12. Yes<br>13. Unclear<br>14. Unclear (no withdrawals) |
| Sach et al (2004) | 15 ALS<br>12 HC | 1.5 | FA | Single-shot stimulated echo acquisition mode (STEAM); not EPI; Diffusion weighting with Stejskal-Tanner spin echo prep<br><br>SPM99; Matlab 5.3 | 6mm FWHM | 6 directions | 750 s/mm2 | Not stated | WM pathways from motor (MC) & premotor cortex (preMC)to brainstem (BS) | • ↓ FA in ALS cf. HC:<br>○ rt PLIC (p < 0.00001)<br>○ lt PLIC (p = 0.012)<br>○ rt.CR under PMC (p = 0.03)<br>○ lt.CR under PMC (p = 0.006)<br>○ rt.CR under PreMC (p = 0.018)<br>○ lt.CR under PreMC (p = 0.001)<br>○ rt pyramidal tract in BS (p = 0.022)<br>○ lt pyramidal tract in BS (p = 0.042)<br>○ CC (p < 0.01)<br>○ rt.Th (p < 0.01) | 1. Yes<br>2. Yes<br>3. Unclear (not all patients fulfilled El Escorial at the time of MRI, but all later fulfilled the El Escorial at follow-up)<br>4. Unclear (see #3)<br>5. Yes<br>6. Yes<br>7. Yes<br>8. Yes<br>9. Yes<br>10. No (El Escorial already known)<br>11. Unclear (6 patients had repeat El Escorial after the MRI scan)<br>12. Yes<br>13. Unclear<br>14. Unclear (no withdrawals) |

| | | | | | | | | | | | |
|---|---|---|---|---|---|---|---|---|---|---|---|
| Borroni et al (2007) | 36 FTD ( 28 fvFTD, 8 tvFTD), 23 HC | 1.5 T | FA | VBM , DTI DTI in single subject using (FACT in Brain Visa software) | 10 mm | 6 | 1000 $s/mm^2$ | NA | SLF, ILF, IFOF | VBM analysis of GM and WM<br>• fvFTD vs HC:<br>GM atrophy ($p<0.05$) in:<br>• Dorsolateral FC<br>• ant Cg C<br>• Insula<br>• Superior temporal G<br>• Bilateral Th<br>• tvFTD vs HC<br>GM atrophy ($p<0.05$) in:<br>• Middle inferior TG<br>• Temporal pole<br>• Orbitofrontal G<br>• Sup TG.<br>• Superior FG.<br>•<br>DTI analysis of WM<br>○ fvFTD vs HC ($p<0.05$)<br>• ↓ FA in rt SLF<br>○ tvFTD vs HC ($p<0.05$)<br>• ↓ FA in bilateral ILF and IFOF<br>• ↓ FA in lt CC, lt SLF<br>Correlation analysis of FA and behavioral and neuropsychological examination<br>• No correlation between FA and demographics<br>• In SLF, -ve correlation between FA and FBI A, FBI B and FBI AB.<br>• Personal neglect (eg, lack of personal hygiene, disorganization in planning and organizing complex activity, impulsivity or poor judgment and utilization behavior were related to ↓ FA in SLF.<br>Multiple regression analysis | 1. Yes.<br>2. Yes (Neary & McKhann refs)<br>3. Yes.<br>4. Unclear.<br>5. Yes.<br>6. Yes.<br>7. Yes.<br>8. Yes.<br>9. Yes.<br>10. No (El Escorial known)<br>11. Yes.<br>12. Yes.<br>13. Unclear.<br>14. Unclear |

| | | | | | | | | | | |
|---|---|---|---|---|---|---|---|---|---|---|
| | | | | | | | | | | • Trail-making Test B scoring have –ve correlation with FA in SLF.<br>• tvFTD correlation were not investigated due to sample size.<br>• The significantly associated FBI sub items independently related to ↓ FA. |

| | | | | | | | | | | | |
|---|---|---|---|---|---|---|---|---|---|---|---|
| Mitsumoto et al (2007) | 43 ALS<br>9 PMA<br>6 PLS<br>6 fALS<br>29 HC | 1.5 | Mean FA, Diffusion constant | SSEPI | NA | 26 | NA | NA | PLIC Pyramidal tract, preCG, preMC | • At Baseline FA and MD (n=64):<br>o No sig in ALS at PLIC<br>o Sig in fALS ($p<0.002$)<br>• Correlation with clinical measures:<br>o No sig with FA or MD with (DD, ALSFRS-R, MMT)<br>• Analysis of changes overtime (n=30):<br>o No sig. changes in imaging markers (from DTI and MRSI)<br><br>Refer to the original paper for more details of other irrelevant results to current review. | 1- Yes<br>2- Yes<br>3- Yes<br>4- Yes<br>5- Yes<br>6- Yes<br>7- Yes<br>8- No<br>9- Yes<br>10- Yes<br>11- Yes<br>12- Yes<br>13- Yes<br>14- Yes |
| Iwata et al (2008) | 31 ALS<br>31 HC | 1.5 | FA | SSEPI<br>ROI | NA | 13 | 1000 s/mm3 | | CST (CR, IC, CP, basis pontis, medulla oblongata) Extramotor WM (gCC, sCC, superior CP, middle CP, inferior CP, cerebellar WM) | • ↓FA in ALS than HC:<br>o Sig at CR, PLIC, CP, basis pontis and medulla oblongata ($p<0.0001$)<br>o Not sig at the extramotor WM<br>• Correlations with clinical measure:<br>o Reduced FA with higher UMN scores at CR, IC, pyramids of medulla oblongata.<br>o Not sig at extramotor WM<br>o No sig with ALSFRS-R in any ROI<br>• Correlation between FA and CMTCs:<br>o Decreased FA along CST with delayed CMCTs and CTX-BS<br><br>more detailed results on TMS can be found in the original paper. | 1. Yes<br>2. Yes (only based on El Escorial criteria without exclusion criteria)<br>3. Yes<br>4. Unclear<br>5. Yes<br>6. Yes<br>7. Yes<br>8. No (no reference to the method or detailed description)<br>9. Yes<br>10. Yes<br>11. Yes<br>12. Yes<br>13. No<br>14. No |
| Matsuo et al (2008) | FTD=20<br>HC=17 | 1.5 | FA, ADC | SS-EPI Tractography (PRIDE)<br><br>ROI | NA | 15 | 1000 s/mm2 | NA | Pyramidal tract (CP), Unci.F,ILF, cc, Arc.F, | mFA measure in FTD and HC:<br>o ↓ FA in association fibers (Arc.F, UnciF, ILF) in FTD than HC ($p<0.01$)<br>o ↓ FA lower in FTD than HC in gCC ($p<0.01$) while slightly decreased in splenium ($p<0.05$).<br>o No sig difference in pyramidal tracts. | 1. Yes<br>2. Yes<br>3. Yes<br>4. Unclear<br>5. Yes<br>6. Yes<br>7. Yes |

| | | | | | | | | | | | |
|---|---|---|---|---|---|---|---|---|---|---|---|
| | | | | | | | | | | ○ No sig difference in between FTD subgroups (early vs advanced) except in lt.UnciF (p<0.05)<br><br>Correlation between FA and neuropsychological scores<br>Sig correlated in lt.UnciF (p<0.05) | 8. Yes<br>9. Yes<br>10. Unclear (diagnosis of FTD already known)<br>11. Unclear<br>12. Yes<br>13. Unclear<br>14. Yes (no withdrawals) |
| Zhang et al (2009) | 18 FTD<br>19 HC | 4 | FA, DR, DA | GRAPPA, Dual SE, EPI<br><br>Volume-one and dTV software (TOI, Voxel-wise Analysis) | FWHM, 4 mm | 6 | 800 s/mm2 | FA = 0.18 | CC, Cg, Unc.F<br>Brain CST | FA:<br>○ Lt.d.Cg, lt.Unc, lt.a.Cg (p<0.0001), rt.Unc (p=0.005), rt.d.Cg (p=0.003), a.CC (p=0.0002), rt.a.Cg (p=0.005)<br><br>RD:<br>○ lt.p.Cg (p=0.008), rt.p.Cg (p=0.01), lt. and rt.a.Cg (p<0.0001), a.CC (p=0.0002), p.CC and rt.CST (p=0.01), rt.d.Cg (p=0.003), L.d.Cg (p=0.001)<br><br>AD:<br>○ a.CC (p<0.0001 ), rt.a.Cg (p= 0.03), L.d.Cg (p= 0.008), R.d.Cg (p= 0.04), L.Unc (p= 0.001), R.Unc (p <0.0001)<br><br>Find results of VWA and TOI in the original paper. | 1. Yes<br>2. Yes<br>3. Yes<br>4. No<br>5. Yes<br>6. Yes<br>7. Yes<br>8. Yes<br>9. Yes<br>10. Yes<br>11. Yes<br>12. Unclear<br>13. Unclear<br>14. Unclear |

| | | | | | | | | | | | |
|---|---|---|---|---|---|---|---|---|---|---|---|
| Chen et al (2009) | 7 FTD<br>20 HC | 1.5 | FA, MD | SSPE-EPI<br><br>ROI analysis (on b=0 then transferred to MD and FA images) | NA | 25 | 1000 s/mm2 | NA | Temporal WM, gCC, sCC, medial SC periventricular area (PV) | • FA<br>○ Lt.Temporal ($p<0.05$) , R.ant.PV ($p<0.05$), bilateral post.PV ($p<0.05$) .<br>• MD:<br>○ Bilateral Temporal ($p<0.05$) , Rt.ant.SC ($p<0.05$), bilateral post.PV ($p<0.05$), gCC ($p<0.05$) | 1. Yes<br>2. Yes<br>3. Yes<br>4. Unclear<br>5. Yes<br>6. Yes<br>7. Yes<br>8. Yes<br>9. Yes<br>10. Yes<br>11. Yes<br>12. Yes<br>13. Unclear<br>14. Unclear |
| Avants et al (2010) | 25 FTD<br>24 AD<br>23 HC | 3 | FA | SSPE-EPI<br><br>Sparse canonical correlation analysis (SCCA) | FWHM, 2 mm | 30 | 1000 s/mm2 | FA>0.2 | CST, ILF, IFOF, Unc.F, CC, SLF | • WM significant correlated with cortical thickness:<br>○ Bilateral CST, ILF, IFOF, UncF, CC and Rmcmillan<br>○ SLF | 1. Yes<br>2. Yes<br>3. Yes<br>4. Unclear<br>5. Yes<br>6. Yes<br>7. Yes<br>8. Yes<br>9. Yes<br>10. Unclear<br>11. Unclear<br>12. Unclear<br>13. Unclear<br>14. Unclear |
| Filippini et al (2010) | 24 ALS<br>24 HC | 3 | FA, MD, AD, RD | Whole brain DWI SE<br>TBSS<br>VBM | NA | 60 | 1000 s/mm$^3$ | NA | CC, CST | FA between ALS vs HC ($p<0.05$ corrected):<br>○ Within CC<br>○ Bilateral WM tracts from central CC to PMC and preMC.<br>○ Rostral CST<br>○ Weaker difference in CST at BS ($p<0.05$ uncorrected)<br><br>↓ FA correlated clinical score in ALS:<br>○ ↑ **UMN score** Bilateral CST ($p<0.05$ corrected) but not in CC<br>○ ↓ **ALSFRS-R** weaker correlation ($p<0.05$ uncorrected) in CC. | 1. Yes<br>2. No<br>3. Yes<br>4. Yes<br>5. Yes<br>6. Yes<br>7. Yes<br>8. Yes (provided online)<br>9. Yes<br>10. No (El Escorial already known)<br>11. Unclear<br>12. Yes |

| | | | | | | | | | | | |
|---|---|---|---|---|---|---|---|---|---|---|---|
| | | | | | | | | | | ○ ↑ **DD** Bilateral CST (p<0.05 corrected)<br><br>↑ RD correlated clinical score in ALS<br>○ In CC and bilateral WM connecting PMC and preMC (p<0.05 corrected)<br><br>No significance found in mean or axial diffusivity.<br><br>↓ GM volumetric atrophy corresponding with FA and RD:<br>○ PMC, preMC, supplementary MC, Cg cortex, TL<br><br>Post hoc combining measure:<br>○ FA correlated with GM differences p= 0.64, $p < 10^{-4}$<br>RD correlated with GM differences GM: p=0.61, $p < 10^{-4}$ | 13. Yes (uncorrected data reported in the result but not shown)<br>14. Yes (no withdrawals) |
| Whitwell et al (2010) | 16 FTD<br>4 SMD<br>7 NFA<br>19 HC | 3 | FA, RD, AD | SSEP FLAIR<br>MPRAGE<br>SENSE<br>(ROI-analysis DtiStudio-software) | FWHM, 8 mm | 21 | NA | 0.05 | CC, Cg, Unc.F, ILF, SLF, CST, AC | • DTI changes in WM in bvFTD (p<0.05 corrected):<br>○ ↓ FA and ↑RD bilateral Unc.F, AC. ant. SLF, ant. ILF with altered RD in gCC and post. ILF<br>○ ↑FA in CST<br><br>Refer to original paper for details of other groups' results and detail FA and RD values. | 1. Yes<br>2. No<br>3. Yes<br>4. Unclear<br>5. Yes<br>6. Yes<br>7. Yes<br>8. Yes<br>9. Yes<br>10. Yes<br>11. Yes<br>12. Yes<br>13. Unclear<br>14. Unclear |
| Agosta et al (2011) | 33 FTLD (bvFTD=13)<br>HCbvFTD=25<br>PPA=20 (NFA=9, semantic | 3 | FA, MD, RD, AD | Pulsed-gradient SE-EPI.<br>WM hyperintensities (WMHs)<br>TBSS.<br>VBM; GM ROI | 8-mm fullwidth at half-maximum (FWHM) | 32 | 1000 s/mm2 | FA=0.2 | WM tracts:<br>CC, Cg, Unci.F, IFOF, ILF, paraHpc, | • **bvFTD vs HC** (p<0.05, corrected**)**<br>○ ↓ FA in CC, Cg, CR, External Capsule (ExC), IC, subcortical WM near frontal cortex (FC), parietal cortex (PC), temporal, occipital WM, Fx, cerebellar peduncle (CP). | 1. Yes<br>2. Unclear (selection criteria clear for HC but not for FTLD)<br>3. Yes<br>4. Unclear<br>5. Yes<br>6. Yes |

| | | | | | | | | | | | |
|---|---|---|---|---|---|---|---|---|---|---|---|
| | =7,<br>logopenic=4)<br>HCPPA=27 | | | | | | | | SLF, CST, Fx<br><br>GM regions:<br>Cg C, FC, preMC, FG, PL, temporal gyri (TG), temporal pole, OC, OFC, white | ○ ↑ MD in CC, Fx, bilateral EC, frontal and temporal WM. Bilateral superior and inferior parietal WM<br>○ ↑ RD similar tracts to the ↓ FA except cerebellum.<br>○ ↑ AD in CC, bilateral orbital and dorsolateral frontal WM, Fx, rtCR, ExC, ant.Temporal WM.<br>• NFA vs HCPPA<br>○ ↓ FA in bilateral (CC, Cg, Fx) but was decreased only in the left hemisphere in (CR, ExC, inferior and dorsolateral frontal WM, temporal and occipital WM.<br>○ ↑ MD in CC, ltCg, ExC, ltCR, lt.orbitofrontal and temporal WM.<br>○ ↑ RD similar to the regions with ↓ FA except the occipital WM.<br>○ ↑ AD in lt.ant dorsolateral frontal WM and lt.inf parietal WM.<br>• Semantic vs HCPPA<br>○ ↓ FA in antCC, lt.CR, ltExc, lt. orbitofrontal WM, lt dorsolateral frontal WM, bilateral ant. Temporal WM<br>○ ↑ AD, RD and MD were found in CC, Cg, CR, ExC and IC, bilateral orbitofrontal and temporal WM, BS and cerebellum<br>• Logopenic vs HCPPA ($p<0.05$ uncorrected)<br>○ ↓ FA in bilateral (CC, Cg, Fx) but was decreased only in the left hemisphere in (CR, ExC, inferior and dorsolateral frontal WM, temporal and occipital WM.<br>○ ↑ MD in CC, ltExC, orbital dorsolateral frontal, inferior pareital and ant.temporal WM.<br>○ ↑ RD in lt. antCC, lt.Cg, lt. orbital and dorsolateral frontal WM, lt inferior parietal WM. | 7. Yes<br>8. Yes<br>9. Yes<br>10. Unclear<br>11. Yes<br>12. Yes<br>13. Yes (in supplementary material)<br>14. Yes (no withdrawals) |

| | | | | | | | | | | | |
|---|---|---|---|---|---|---|---|---|---|---|---|
| | | | | | | | | | | ○ ↓ FA in lt.ant CC, lt. ant and post. Cg, ltCR, lt Exc, bilateral orbitofrontal and left inferior frontal WM, lt temporal and lt. inferior parietal WM.<br>○ ↑ AD in CC, lt.Cg, Fx, lt Temporal and inferior parietal WM | |
| Hornberger et al (2011) | 14 bvFTD<br>15 AD<br>18 HC | 3 | FA, | TBSS | FWHM, 8 mm | 32 | 1000 s/mm2 | NA | Unc.F, CF, FMF, Cg, FM, CC | • **DTI ↓ FA** ($p<0.001$, uncorrected) **in:**<br>○ Unc.F<br>○ CF<br>○ FMF<br>• VBM ↓GM in bvFTD vs HC<br>○ a.Cg<br>○ OF<br>○ Insula<br>○ a.TL<br>• Correlation:<br>○ FA of UncF and Hayling task, -ve.<br>○ FA of FM and Cg<br>○ FA of UncF and Neuropsychiatric inventory disinhibition= -ve | 1. Yes<br>2. Yes<br>3. Yes<br>4. Unclear<br>5. Yes<br>6. Yes<br>7. Yes<br>8. Yes<br>9. Yes<br>10. Yes<br>11. Unclear<br>12. Yes<br>13. Unclear<br>14. Unclear |
| Sarro et al (2011) | 16 ALS<br>15 HC | 1.5 | FA, MD, RD, AD | FLAIR<br>Pulsed-gradient EPI<br>Tractography,<br>Fmri | NA | 12 | 1000 s/mm3 | 0.4 | CC, CST, Cg, SLF, IFOF, ILF, UnciF, Fx | • FLAIR hyperintensities:<br>○ Bilateral CST; caudal PLIC, CR and ventral BS<br>• DTI ALS vs HC:<br>○ ↑ MD of CC ($p<0.04$), CST (right, $p=0.001$, left, $p=0.002$), bilateral SLF ($p=0.02$), rt.Cg ($p=0.2$), bilateral Unci.F ($p=0.02$)<br>○ ↓ FA in CST (right, $p=0.001$, left, $p=0.002$)<br>○ ↑ RD in bilateral CST ($p<0.001$), bilateral SLF ($p<0.03$), Unci.F ($p=0,3$)<br>○ ↑ AD in rt.SLF ($p=0.4$), rt.Cg ($p=0.3$), bilateral Unci.F (right, $p=0.03$, left, $p=0.002$)<br>• Regression analysis between DTI metrics and neuropsychological tests:<br>○ Trail-making test scores correlated with CC (MD and FA), rt. ILF (MD), CST, | 1. YES<br>2. YES<br>3. YES<br>4. YES<br>5. YES<br>6. YES<br>7. YES<br>8. YES<br>9. YES<br>10. YES<br>11. YES<br>12. YES<br>13. Unclear (reported only significant data)<br>14. Yes (excluded 2 patients) |

| | | | | | | | | | | | |
|---|---|---|---|---|---|---|---|---|---|---|---|
| | | | | | | | | | | Cg, IFOF, ILF all bilaterally (FA), rt. Unci.F (FA).<br>○ Stroop test scores associated with CC, rt.IFOF, ILF (FA)<br>○ Wisconsin Card sorting test correlated with bilateral IFOF and ILF (MD), rt.IFOF (FA).<br>○ Phonemic fluency test scores correlated with lt. Cg (FA)<br>○ Verbal learning and memory test scores correlated with Fx (FA and MD)<br>○ Visual-spatial correlated with Unci.F (FA) | |
| Tsujimoto et al (2011) | 21 ALS<br>21 HC | 3.0 | FA<br>MD | Stejskal-Tanner sequence with Single-shot SE EPI<br><br>dTV-II.SR; Volume-One 1.72; SMP5 | 8mm isotropic Gaussian kernel | Not stated | 1000 s/mm2 | Cluster extent threshold = 20 voxels | CST; Frontal lobe areas | • ↓ FA in bilateral CST and frontal lobe gyri; ↑ MD in bilateral CST (all $p < 0.001$; visual no absolute values)<br>• Correlations between FA and increased apathy score on FrSBe:<br>○ Rt. mFG, R sub-gyral FL areas, rt. middle FG, R para.Hpc G, rt.ant.CG, lt Th, lt.sup.FG (all $p \leq 0.002$)<br>• Correlations between MD and increased apathy score in FrSBe:<br>○ Lt.sup FG, rt.sup.TG (all $p \leq 0.002$); see tables in paper | 1. Yes<br>2. Yes<br>3. Yes<br>4. Yes<br>5. Yes<br>6. Yes<br>7. Yes<br>8. Yes<br>9. Yes<br>10. No (El Escorial already known)<br>11. Yes<br>12. Yes<br>13. Unclear<br>14. Unclear (exclusions not withdrawals) |
| Zhang et al (2011) | 20 FTD<br>20 AD<br>21 HC | 4 | FA | dual spin-echo EPI, GRAPPA (dTV.II software) | FWHM, 8 mm | 6 | 800 s/mm2 | NA | FL, TL CC, Cg Unc, Th, Caudate, LPL. | • DTI ↓ FA in FTD vs HC ($p<0.001$, uncorrected):<br>○ frontal and temporal lobes<br>○ a.CC<br>○ a.Cg<br>• VBM ↓GM in bvFTD vs HC<br>○ Frontal and temporal lobes<br>○ Rt. frontoinsular gyrus<br>○ Limbic lobes such as bilateral anterior cingulate gyrus<br>○ Uncus | 1. Yes<br>2. Yes<br>3. Yes<br>4. Unclear<br>5. Yes<br>6. Yes<br>7. Yes<br>8. Yes<br>9. Yes<br>10. Unclear<br>11. Yes<br>12. Yes |

| | | | | | | | | | | | |
|---|---|---|---|---|---|---|---|---|---|---|---|
| | | | | | | | | | | ○ Subcortical nuclei including the bilateral caudate<br>○ The thalamus<br>○ The lateral parietal lobes. | 13. Unclear<br>14. Unclear |
| McMillan (2012) | 50 FTLD<br>42 AD<br>38 HC | 3 | FA | SSPE-EPI, MPRAGE (TSA) (whole-brain GM density analyses.) | FWHM, 4 mm | 30 | 1000 s/mm2 | NA | CC, CST, IFOF, ILF, SLF, UncF | • DTI ↓ FA in FTD vs HC<br>○ bilateral CST, IFO, ILF, SLF, and UncF, CC most prominent in anterior portions of these tracts.<br><br>• DTI ↓ FA in FTLD vs AD<br>○ More in the ant.CC in FTLD<br>○ No sig areas in AD group<br><br>Other irrelevant results on GM density are on the research paper. | 1. Yes<br>2. Yes<br>3. Yes<br>4. Yes<br>5. Yes<br>6. Yes<br>7. No<br>8. Yes<br>9. Yes<br>10. No (El Escorial already known)<br>11. Yes<br>12. Yes<br>13. Yes (provided online)<br>14. Yes |
| Lillo et al (2012) | 10 ALS,<br>10 ALS-FTD,<br>15 bvFTD,<br>18 HC | 3.0 | FA | FDT toolbox in FSL; TBSS | Not stated for DTI | 32 gradients | 1000 s/mm2 | Unclear | Whole brain | • Comparisons: ($p < 0.05$ FWE corrected)<br>○ **bvFTD vs. HC:** WM degeneration in FM, ant. CC, ant. ILF, CST<br>○ **ALS-FTD vs. HC:** WM degeneration same as above, but less in FM and ant. CC (more in ant ILF and CST)<br>○ **ALS vs. HC:** WM degeneration in CST (only small changes in FM, ant. CC and ILF)<br>○ **bvFTD vs. [ALS-FTD and ALS]:** more WM degeneration in FM, ant. CC and ILF; less in CST and temporal poles<br>○ **ALS-FTD vs. ALS:** more WM degeneration in FM, ant. CC and ILF; less in CST | 1. Yes<br>2. Yes (revised FTD Criteria in 2011 Brain paper)<br>3. Yes<br>4. Yes<br>5. Yes<br>6. Yes<br>7. Yes<br>8. Yes<br>9. Yes<br>10. No (ALD/FTD classification already known at time of MRI)<br>11. Yes<br>12. Yes<br>13. Unclear<br>14. Unclear (no withdrawals) |

| | | | | | | | | | | | |
|---|---|---|---|---|---|---|---|---|---|---|---|
| Pettit et al (2012) | 30 ALS<br>30 HC | 1.5 | FA, MD | Whole brain Single shot SE EPI | NA | 64 | 1000 s/mm3 | 0.2-0.4 | Cg, Th, SFG, CR, CC, UnciF, CST, ILF, SLF, superior PL, temporal gyri, OG, optic radiation. | ROI analysis sig. gp difference in frontal regions:<br>○ Ant. Cg, ant.ThR (FA,MD) Unci.F (MD)<br>In prefrontal regions:<br>○ Sup., inf., mid FG (FA, MD)<br>○ Ant.CR (MD)<br>In temporal regions:<br>○ hippocampus and Cg (MD)<br>○ Temporal G, ILF, CR (FA)<br>CST and CC (FA, MD)<br>CC segmentation:<br>○ FA (P=0.02)<br>○ MD (P=0.02)<br>No sig between DTI measures and optic radiation or OG<br>No sig correlation with FVC, onset site or cognitive tests<br>Neuropsychological data correlation with MRI<br>Letter fluency:<br>○ Sup.FG (MD, p=0.012),<br>○ inf.FG (FA, p=0.017),<br>○ CST (FA, p=0.01),<br>○ CC (FA, p=0.049)<br>Reverse digit span<br>• hippocampus of Cg (MD, p=0.018) | 1. Yes<br>2. Yes<br>3. Yes<br>4. Yes (disease duration presented and consistent)<br>5. Yes<br>6. Yes<br>7. Yes<br>8. Yes<br>9. Yes<br>10. No (El Escorial already known)<br>11. Yes<br>12. Yes<br>13. Yes (table containing all significant and non-significant results)<br>14. Unclear |
| Prell et al (2012) | 17 ALS<br>17 HC | 1.5 | FA<br>ADC | EPI<br><br>SPM2;<br>MATLAB 12 | 6mm FWHM | 24 directions | 1000 s/mm2 | Unclear | Whole brain | • Multiple areas of significance (large tables and images in paper); no raw values given, but set at p < 0.05 corrected.<br>• Comparisons:<br>○ ADC in ALS vs. HC<br>○ FA in ALS vs. HC<br>○ ADC in Bulbar ALS vs. HC<br>○ ADC in Limb ALS vs. HC<br>○ FA in Bulbar ALS vs. HC<br>○ FA in Limb ALS vs. HC<br>○ ADC in Bulbar ALS vs. Limb ALS (n.s.) | 1. Yes<br>2. Yes (El Escorial , but no ref given)<br>3. Yes<br>4. Unclear (not stated which El Escorial category they were at the time of the scan)<br>5. Yes<br>6. Yes<br>7. Yes<br>8. Yes |

| | | | | | | | | | | | |
|---|---|---|---|---|---|---|---|---|---|---|---|
| | | | | | | | | | | ○ FA in Bulbar ALS vs. Limb ALS (n.s.) | 9. Unclear (no ref provided)<br>10. No (El Escorial already known)<br>11. Yes<br>12. Yes<br>13. Unclear<br>14. Yes |
| Prudlo et al (2012) | 15 ALS<br>7 LMN variant<br>21 HC | 1.5 | FA | EPI<br><br>Whole-brain TBSS (using FSL 4.1); followed by ROI analysis of selected WM regions (DTIstudio version 2.4.01) | States TBSS does not need a smoothing kernel | 30 directions | 1000 s/mm2 | Not stated | Whole brain | [Comparisons at $p < 0.05$ significance level]<br>↓ FA in All ALS (n=22) vs. HC (n=21):<br>○ CST, ATR, aLIC, pLIC, GCC, Fj, FM, UncF, SLF, ILF, IFOF, Cg, MCP<br>↓ FA in Classic ALS (n=7) vs. HC (n=7):<br>○ CST, ATR, aLIC, pLIC, bCC, Fj, FM, Unc.F, SLF, ILF, IFOF, Cg, MCP, ICP<br>↓ FA in LMN -ALS (n=7) vs. HC (n=7):<br>○ CST, ATR, aLIC, pLIC, GCC, Fj, FM, UF, SLF, ILF, IFOF, MCP, ICP<br>↓ FA in Classic ALS (7) vs. LMN variant (7):<br>○ No sig | 1. Yes<br>2. Yes<br>3. Yes<br>4. Yes<br>5. Yes<br>6. Yes<br>7. Yes<br>8. Yes<br>9. Yes<br>10. No (El Escorial already known)<br>11. Yes<br>12. Yes<br>13. Unclear<br>14. Unclear (no withdrawals) |
| Bastin et al (2013) | 30 ALS<br>30 HC | 1.5 | FA, MD,RD | Whole brain SE EPI<br><br>TractoR (tract segmentation) | | 64 | 1000 s/mm3 | | Genu and splenium CC, CCG, CST, UnciF, ILF, Arcuate F | Tract integrity ALS vs HC<br>○ ↑ MD in all 12 tracts and significant in rt. CCG (p=0.03), lt.CST (p=0.02), rt.CST (p=0.001),<br>○ ↓ FA in lt. CST(p=0.002), rt. CST (P=6X10-5), rt. UnciF (p=0.07)<br><br>Tract shape difference:<br>○ Sig. in lt.CST (p=0.04), rt. CST (p=0.02), rt. UnciF ($p<0.05$)<br>Correlation with disease progression:<br>○ ⇓ correlation with FA along lt.CST (p=0.02), rt. CST (p=0.01)<br>○ ⇓ correlation with RD along lt.CST (p=0.07) | 1. Yes<br>2. Yes<br>3. Yes<br>4. Yes<br>5. Yes<br>6. Yes<br>7. Yes<br>8. Yes (appears to have enough detail about MRI method)<br>9. Yes<br>10. No (El Escorial already known)<br>11. Unclear<br>12. Yes |

| | | | | | | | | | | | |
|---|---|---|---|---|---|---|---|---|---|---|---|
| | | | | | | | | | | | 13. Yes (table with all significant and non-significant results)<br>14. Yes (same number in the result section) |
| Bede et al (2013) | 39 ALS (C9pos=9, C9neg=30)<br>44 HC | 3 | FA, MD, RD | SE-EPI<br>TBSS,<br>VBM,<br>Cortical thickness. | NA | 32 | 0,1,100 s/mm2 | NA | CC, CST, cerebellar pathways, superior motor tracts | • DTI in C9pos vs C9neg ($p<0.05$, corrected):<br>○ FA, RD,MD changes in frontotemporal abnormalities in C9pos<br>○ More changes involved in motor WM.<br>○ Overlap WM pathology in both groups was significant in bCC and superior CST.<br>○ gCC, AC and bilateral Th related to genotype-specific gp.<br>• Cortical thickness analysis<br>○ Atrophy GM in C9pos vs C9neg: lt.fusiform, lt. supramarginal, lt.sup TG, lt. OFC, lt. lateral OC, lt. posterior CG.<br>○ C9pos vs HC: bilateral prefrontal cotices, insular cortex, fusiform G, supramarginal C, lateral OC, precuneus, temporal poles, inferior FG.<br>○ No significant findings in C9neg and HC<br>• VBM:<br>○ Orbitofrontal, opercular and temporal changes in the C9pos compared to HC and C9neg | 1. Yes<br>2. Yes<br>3. Yes<br>4. Yes<br>5. Yes<br>6. Yes<br>7. Yes<br>8. Yes<br>9. Yes<br>10. Unclear<br>11. Unclear<br>12. Yes<br>13. Unclear<br>14. Yes (no withdrawals) |
| Bozzali et al (2013) | FTLD(+GRN) =6<br>FTLD(-GRN) =17<br>HC=12 | 1.5 | FA, MD | DW SE-EPI<br>VBM<br>Probabilistic tractography | 10mm FWHM Gaussian kernel | 12 | 1000 s/mm2 | probabilistic threshold=0.1 + n 0.02 | CC divided to 5 regions:<br>1.Most anterior - prefrontal cortex (PFC)<br>2.Rest of ant CC-PMC and | DTI between FTLD and HC:<br>○ Widespread pattern of ↓ FA and ↑ MD across all regions of CC.<br><br>Similar finding when comparing each group against HC.<br><br>DTI between FTLD(+GRN) and FTLD(-GRN)<br>○ ↓ FA and ↑ MD in the most antCC; bilaterally more prominent in the left side. | 1. Yes<br>2. No (same subjects recruited for previous study)<br>3. Yes<br>4. Yes<br>5. Yes<br>6. Yes<br>7. Yes<br>8. Yes<br>9. Yes<br>10. Yes |

| | | | | | | | | | | | |
|---|---|---|---|---|---|---|---|---|---|---|---|
| | | | | | | | | | supplment ary MC.<br>3.mid CC-pre-rolandic C.<br>4.posterior CC-post-rolandic cotices.<br>5.most posterior CC-connecting post. Parietal, temporal, occipital cortices. | VBM between FTLD(+GRN) and FTLD(-GRN)<br>○ ↓ mGM near the most antCC<br>Correlation analysis<br>○ VBM: mGM positively correlated with mean FA and negatively with mean MD for the most antCC in FTLD(+GRN) but not FTLD(-GRN)<br>○ Voxel wise analysis (VWA) confirmed the same findings as VBM between mGM and mean FA particularly in lt.CC. | 11. Yes<br>12. Yes<br>13. No<br>14. Yes (no withdrawals) |
| Furtula et al (2013) | 30 HC; 14 ALS | 3T | FA | Double spin echo single shot EPI<br><br>Deterministic tractography | | 26 | 1000 s/mm2 | | CST, PLIC | • No significant difference in mean FA between ALS and HC (CST, PLIC)<br><br>• In ALS, no significant difference in FA between more affected and less affected side of CST<br><br>• No correlation between FA (CST or PLIC) and ALSFRS-R or disease duration | 1. Unclear (2 patients diagnosed with PMA)<br>2. Yes (ALS according to El Escorial + 2 extra PMA patient recruited)<br>3. Yes<br>4. Yes<br>5. Yes<br>6. Yes<br>7. Yes<br>8. Yes<br>9. Yes<br>10. No (El Escorial / PMA status already known)<br>11. Yes<br>12. Yes<br>13. Yes (all data completely reported for each individual patient)<br>14. Yes (withdrawal(s) explained) |

| | | | | | | | | | | | |
|---|---|---|---|---|---|---|---|---|---|---|---|
| Frizell Santillo et al (2013) | 14 bvFTD<br>22HC | 3 T | FA,MD, RD, AD | Track Vis Tractography QuTE analysis | NA | 48 | 800 s/mm$^2$ | 0.2 | Cg | DTI bvFTD vs HC:<br>• ↓ FA in the ant Cg.but not the posterior part.<br>• Lt hemisphere is slightly more affected more than rt.<br>• ↓ FA (p<0.001) and ↓ MD (p<0.003) and ↑ RD (p<0.002) and ↑ AD (p<0.002) but the RD was the DTI that showed greatest differences between bvFTD and HC followed by MD.<br><br>VBM bvFTD vs HC:<br>• Moderate differences in the lt more than the rt but not statistically significant.<br>• 4 possible bvFTD showed slightly significant cortical thinning compared to HC (p=0.022).<br><br>Correlation between DTI and VBM:<br>• In the lt hemisphere VBM correlated with FA (p=0.002) and MD (p<0.001), RD (p<0.001) and AD (p=0.001).<br><br>Regression analysis between DTI and cortical integrity:<br>• AUC of VBM and DTI parameters were larger in the lt hemisphere than the rt.<br><br>Thickness can correctly classify 78% of cases, VBM 83%, FA 84%, MD 90%, AD 88% and RD 92%. | 1. Yes.<br>2. Yes (part of Lund Prospective FTD study)<br>3. Yes.<br>4. Yes (mean +/- SD provided)<br>5. Yes.<br>6. Yes.<br>7. Yes.<br>8. Yes.<br>9. Yes (Rascovsky reference provided)<br>10. No (El Escorial known)<br>11. Yes.<br>12. Yes.<br>13. Unclear.<br>14. Unclear |
| Grapperon et al (2013) | 14 ALS<br>6 LMN syndrom e<br>13 HC | 1.5 | FA, MD, RD,AD | SE-EPI, T2, FLAIR Tractography | NA | 30 | 1000 s/mm3 | 0.2 | CST | Structural CST impairment in ALS gp:<br>○ ↓ Z-MD (p=0.004) and Z-RD (p=0.001)<br>○ ↑ Z-FA (p=0.055).<br><br>Correlation DTI vs clinical parameter:<br>○ Z-MD of lt.CST correlated with UMN scores (p=0.014) and Z-RD of lt.CST (p=0.048)<br><br>No correlation with ALSFRS-R, DD, disease progression | 1. Yes<br>2. Yes<br>3. Yes<br>4. Yes (2 years recruitment)<br>5. Yes<br>6. Yes<br>7. Yes<br>8. Yes<br>9. Yes<br>10. No (El Escorial already known) |

| | | | | | | | | | | | |
|---|---|---|---|---|---|---|---|---|---|---|---|
| | | | | | | | | | | (Unrelated results not reported) | 11. Yes (El Escorial applied before MRI was done)<br>12. Yes<br>13. Unclear<br>14. Unclear (no withdrawal) |
| Rajagopalan et al (2013) | 12 HC; 87 ALS (4 subgroups) | 1.5 T | FA, MD, AD, RD | Single-shot EPI Fiber tracking (FACT)<br>ROI analysis | Not stated | 12 | 1000 s/mm2 | 0.2 | CST at four levels:<br>1. cerebral peduncle (CP)<br>2. PLIC<br>3. Centrum semiovale at top of lateral ventricle (CSoLV)<br>4. subPMC | • 4 subgroups:<br>o UMN-CST hyperintense+<br>o UMN-CST hyperintense-<br>o Mixed UMN/LMN<br>o ALS-FTD<br>• ALL ALS vs. controls:<br>o ↓ FA at IC, CSoLV, subPMC (not CP)<br>o ↓ FA at left subPMC only in UMN groups<br>o ↓ FA at right PLIC in all 4 ALS groups<br>o MD in lt and rt PLIC and lt. CSoLV in ALS-FTD, compared with both HC and other ALS groups<br>o ↑ RD at lt.CP in all ALS groups<br>o ↑ RD at lt. IC, and rt and lt.CSoLV in ALS-FTD<br>• Correlation with clinical measures when all ALS patient pooled together, no FDR correction<br>o MD and RD versus ALSFRS-R scores=-ve (p=0.001) at lt. CSoLV<br>o AD and Disease duration=+ve (p=0.001) at lt CSoLV.<br>o FA and disease progression rate=-ve at IC; rt (p=0.002) and lt. (p=0.001)<br>o FA and disease progression rate=-ve at lt and rt CSoLV (p=0.001 and p=0.018)<br>• No significant correlations when correction for multiple comparison | 1. Yes<br>2. Incomplete (controls not specified appropriately)<br>3. Yes<br>4. Yes<br>5. Yes<br>6. Yes<br>7. Yes<br>8. Yes<br>9. Yes<br>10. No (El Escorial already known)<br>11. Yes<br>12. Yes<br>13. Yes<br>14. Unclear<br><br>** Note this study's presentation of data has some errors (e.g. comparing what is reported in the text and in the figures) |

| | | | | | | | | | | | |
|---|---|---|---|---|---|---|---|---|---|---|---|
| Trojsi et al (2013) | 19 ALS,<br>19 HC | 3T | GFA | GRE EPI<br>TBSS<br>VOI | NA | 32 | 1000 s/mm3 | >0.1 | CST, CC, SLF,Unc.F,<br>FOF | ↓ GFA ALS vs HC<br>○ CST (p<0,05, corrected) within rostral WM below lt and rt.pre.CG, lt.PMJ, ant.Th, ant.Cg, splenium and body CC, Fx, lt. UncF, lt.ant.FOF, lt.SLF<br><br>Correlation ALS vs clinical measures<br>○ ⇓ with UMN scores: along lt. CST at PMJ (p=0.031)<br>○ ⇓ with UMN scores: body CC, ant.Cg,SLF and bi.WM tracts from Central CC to pri.MC and preMC.<br>○ ⇓ with FrSBe: WM in lt. priMC, Body CC, lt.SLF, CST at PMJ<br>○ No sig between GFA and ALSFRS-R, DD, Disease progression in above tracts. | 1. Yes<br>2. Yes<br>3. Yes<br>4. Yes (disease duration presented and consistent)<br>5. Yes<br>6. Yes<br>7. Yes<br>8. Yes<br>9. Yes<br>10. No (El Escorial already known)<br>11. Unclear<br>12. Yes<br>13. Unclear<br>14. Unclear (same number of pt.) |
| Zhang et al (2013) | 13 bvFTD,<br>6 SD, 6 PNFA,<br>19 HC | 4.0 | FA<br>AD<br>RD | Twice-refocused spin-echo diffusion EPI, supplemented with GRAPPA<br><br>SPM8; R; dTV; Volume-one | 4mm FWHM Gaussian kernel | 6 non-collinear directions | 800 s/mm2 | FA = 0.2 | Whole brain, followed by specific TOI (ant CC, post CC, Cg, paraHpc, uncF, AF, Fx) | • Overall: DTI (especially RD) most accurate for FTD subtype classification<br>• FA, RD and AD all presented visually in paper (in each subtype of FTLD vs. HC)<br><br>TOI analysis bvFTD vs HC (supplementary data online)<br>• ↑RD in rt.paraHpc (p<0.02), Fx, lt a.Cg, bilateral UncF,a.CC and p.CC (p<0.001), lt.paraHpc (p<0.04), rt.paraHpc (p<0.03),<br>• ↓FA in the Fx, a.CC, lt.aCg (p<0.001), p.CC (p<0.03), lt.UncF (p<0.005), and rt.UncF (p<0.002).<br>• ↑AD in the aCC, Fx, and bilateral UncF (p<0.001), pCC(p<0.03), and rt.paraHpc (p<0.05). | 1. Yes<br>2. Yes (Neary criteria reference)<br>3. Yes<br>4. Yes<br>5. Yes<br>6. Yes<br>7. Yes<br>8. Yes<br>9. Yes<br>10. No (classification already known when analysing MRI data)<br>11. Yes<br>12. Yes<br>13. …<br>14. Unclear (no withdrawals) |

| | | | | | | | | | | | |
|---|---|---|---|---|---|---|---|---|---|---|---|
| Agosta et al (2013) | 35 HC;<br>26 PLS;<br>28 ALS | 3T | FA<br>MD<br>AD<br>RD | T2-weighted<br>SE<br>FLAIR<br>T1 FFE<br>Pulsed-gradient SE echo planar<br>TBSS<br>Probabilistic tractography<br>ROI analysis | Not stated | 32 | 1000 s/mm2 | 0.2 | CST<br>CC | • PLS vs. HC (p < 0.05, FWE):<br>o ↓FA in entire CST (pyramids – PLIC -preCG), CC (midbody, genu, splenium), ALIC, SLF, Fx, Th, parietal lobes<br>o ↑MD and RD in CST, CC (midbody), ALIC, thalamus, parietal lobes<br>• ALS vs. HC (p < 0.05, FWE):<br>o ↓FA (and increased RD) in entire CST (pyramids –PLIC – preCG,CR), CC (midbody), SLF, parietal lobes<br>o No significant differences in MD<br>• PLS vs. ALS (p < 0.05, FWE)<br>o ↓FA in CC (mid-body), patchy areas of motor, premotor, prefrontal and parietal WM, cerebellum<br>o ↑MD and ↑RD in CC (mid-body), cerebral peduncles, thalamic radiations, Fx, rt. PLIC, patchy areas of motor and premotor WM<br>• TRACTOGRAPHY:<br>o PLS vs. HC:<br>▪ CST, CC-PMC, CC-SMA<br>o ALS vs. HC:<br>▪ CST, CC-PMC<br>o PLS vs. ALS:<br>▪ CC-PMC, CC-SMA | 1. Yes<br>2. Yes<br>3. Yes<br>4. Yes<br>5. Yes (El Escorial, or reference for PLS diagnostic criteria)<br>6. Yes<br>7. Yes<br>8. Yes<br>9. Yes<br>10. No (El Escorial / PLS already known)<br>11. Yes<br>12. Yes<br>13. Yes (all non-significant results appear to be reported)<br>14. Unclear (no withdrawals reported) |
| Barbagallo et al (2014) | 24 ALS<br>22 HC | 3T | FA, MD | SE-EPI<br><br>Automated ROI (FSL) for DTI processing FIRST for T1-WI | NA | 27 | 1000 s/mm2 | NA | Caudate, putamen, globus pallidus, Th, hippocam pus, Amygdala (Ag), frontal | • DTI metrics were averaged due to absence of difference between lt vs rt .<br>• DTI in ALS (n=24) vs HC:<br>o ↑ MD in patient at FC (p=0.023), caudate (p=0.01), Th (p= 0.019), Hpc (p=0.002) and Ag (p= 0.012).<br>o No sig in FA in any structure.<br>• Correlation between clinical features and MD:<br>o +ve correlation with DD in caudate (p=0.004), Th (p=0.02), FC (p= 0.01). | 1. Yes<br>2. Yes<br>3. Yes<br>4. Yes<br>5. Yes<br>6. Yes<br>7. Yes<br>8. Yes<br>9. Yes<br>10. Unclear<br>11. Yes |

| | | | | | | | | | | | |
|---|---|---|---|---|---|---|---|---|---|---|---|
| | | | | | | | | | vortex (FC) | ○ –ve correlation with ALSFRS-R scores in Th (p=0.02), Ag (p=0.02) and FC (p=0.001).<br>• Correlation between neuropsychological scores (n=13) and MD:<br>○ -ve correlation with MSCT in caudate (p=0.007), hippocampus (p=0.001),Ag (p=0.015) and FC (p= 0.002).<br>○ -ve correlation with FAB scores caudate (p=0.017), hippocampus (p=0.001), Ag (p= 0.002) and FC (p=0.009). | 12. Yes<br>13. Yes<br>14. Yes |
| Cardenas-Blanco et al (2014) | 29 HC; 28 ALS (14 limb, 14 bulbar); FTD specifically excluded | 3T | FA, MD, RD, axial diffusivity (AD) | Single-shot EPI GRAPPA TBSS (whole-brain)<br>ROI analysis | Not stated | 30 | 1000 s/mm2 | FA >0.2 | CST | • ALS-Bulbar (TBSS):<br>○ ↓FA in CST ($p < 0.05$ corrected);<br>○ RD ↑only at thalamic CST;<br>○ no changes in MD or AD<br>• ALS-Limb (TBSS):<br>○ patchy ↓FA and ↑RD in CST (only at $p < 0.05$ uncorrected)<br>○ no changes in MD or AD<br>• ROI Analysis:<br>○ also showed significantly ↑MD in ALS-Bulbar CST<br>○ significantly ↓AD in ALS-Limb CST | 1. Yes<br>2. Yes<br>3. Yes<br>4. Unclear time-frame<br>5. Yes<br>6. Yes<br>7. Yes<br>8. Yes<br>9. Yes<br>10. No (El Escorial already known)<br>11. Yes<br>12. Yes<br>13. Unclear<br>14. Unclear (no withdrawals) |
| Christidi et al (2014) | 21 ALS (not FTD); 11 HC | 3T | FA<br>ADC<br>AD<br>RD | Single-shot spin-echo EPI<br><br>Quantitative tractography | NA | 30 | 1000 | 0.15 | Bilateral UncF | ALS vs. HC:<br>• ↑ AD in the bilateral UncF in ALS, compared with HC ($p < 0.05$); other measures not significant | 1. Yes.<br>2. Yes.<br>3. Yes.<br>4. Yes (mean +/- SD provided)<br>5. Yes.<br>6. Yes.<br>7. Yes.<br>8. Yes.<br>9. Yes<br>10. No (El Escorial known)<br>11. Yes.<br>12. Yes.<br>13. Unclear. |

| | | | | | | | | | | | |
|---|---|---|---|---|---|---|---|---|---|---|---|
| | | | | | | | | | | | 14. Unclear (no withdrawals) |
| Crespi et al (2014) | 20 HC; 19 non-demented ALS (a subset of the whole study cohort) | 3T | FA<br>MD<br>Mode of anisotropy (MO) | Single-shot EPI<br><br>TBSS<br><br>Probabilistic tractography | Gaussian kernel (3mm) | 32 | 1000 s/mm2 | 0.2 | CST, SLF, ILF, IFOF, CC and other commissural fibers | • ALS vs. HC:<br>○ ↓FA ($p < 0.05$ FWE) in bilateral CST, body of CC<br>○ No significant differences in MD<br>○ Abnormal MO ($p < 0.05$ FWE) in right CST and ($p < 0.005$ uncorrected) in SLF, IFOF, ILF, genu of corpus callosum and forceps minor<br><br>• Correlation between FA/MO of rt. ILF and IFOF, and emotional recognition (faces) in ALS<br>○ +ve global performance and mean FA in rt.ILF (p=0.04)<br>○ +ve relationship with cumulative scores of assessing negative emotions and the ability to recognize specific negative emotions with FA in rt.ILF (p=0.004) specifically with identification of fear (p=0.03), disgust (p=.0.01) and sadness (p=0.008) while in IFOF (p=0.02) correlated with fear (p=0.008), anger (p=0.05) and sadness (p=0.02)<br>○ No correlation between emotion recognition abilities and mean MO along tracts of interest.<br><br>For additional analyses refer to the result section of the paper. | 1. Yes<br>2. Yes<br>3. Yes<br>4. Yes (durations clearly specified)<br>5. Yes<br>6. Yes<br>7. Yes<br>8. Yes<br>9. Yes<br>10. No (El Escorial already known)<br>11. Yes<br>12. Yes<br>13. Yes<br>14. Yes (withdrawals from MRI stage clearly explained) |

| | | | | | | | | | | | |
|---|---|---|---|---|---|---|---|---|---|---|---|
| Floeter et al (2014) | 28 HC; 25 PLS, 22 ALS | 3T | FA<br>MD | Single-shot EPI<br><br>TBSS | Not stated | 32 (Phillips) or 80 (GE) | 1000 s/mm2 (Phillips) or 300 / 1100 s/mm2 (GE) | Not stated | Subcortical WM, middle cerebellar peduncles (MCP), pons, PLIC, ALIC, thalamus, CC | • All patients (ALS/PLS) vs. HC:<br>o widespread decreased FA and ↑MD (subcortical WM, IC, CC) P<0.05 corrected<br><br>• Patients with Pseudobulbar Affect (PBA+) vs. those without (PBA-) – ALS and PLS introduced as covariates:<br>o reduced FA at left sub-preCG WM; Increased MD at bilateral MCP, transverse pontine fibers, bilateral PLIC, lt. ALIC, Th, frontotemporal subcortical WM, bilateral sub-preCG WM, and CC ( P<0.05 corrected) | 1. Yes<br>2. Yes<br>3. Yes<br>4. Yes (retrospective, so all diagnoses confirmed)<br>5. Yes<br>6. Yes<br>7. Yes<br>8. Yes<br>9. Yes<br>10. No (El Escorial already known)<br>11. Yes<br>12. Yes<br>13. Yes<br>14. Unclear (no withdrawals – retrospective) |
| Heimrath et al (2014) | 11 HC; 9 ALS | 3T | FA | Resting-state fMRI (EPI), with functional connectivity analysis Tensor imaging and fiber tracking (TIFT) | 8mm FWHM (for fMRI) ; 6mm kernel (for DTI) | 12 | 800 s/mm2 | 0.2 | medial prefrontal cortex (MPFC), inferior PL (IPL), Posterior Cingulate cortex (PCC), Para hippocampus (paraHpc)<br><br>Tracts to: anterior PC, orbitofrontal cortex (OFC), PreCG, superior PL, SMG, | • ALS: ↑connectivity in resting (default-mode network) between:<br>o Lt IPL – Rt IPL<br>o Lt IPL – Lt paraHpc<br>o Rt IPL – Lt paraHpc<br>o Lt paraHpc – PCC<br>• ALS:<br>o significantly ↓FA ($p<0.05$) in tracts to Rt anterior PC, R OFC, PreCG (Rt and Lt), Lt superior PL, Lt SMG, Th. | 1. Yes<br>2. Yes<br>3. Yes<br>4. Yes (time since Dx specified)<br>5. Yes<br>6. Yes<br>7. Yes<br>8. Yes<br>9. Yes<br>10. No (El Escorial already known)<br>11. Yes<br>12. Yes<br>13. …<br>14. Yes (reasons for withdrawals/ post-recruitment exclusions identified) |

| | | | | | | | | | | | |
|---|---|---|---|---|---|---|---|---|---|---|---|
| | | | | | | | | | thalamus (Th) | | |
| Irish et al (2014) | 11 bvFTD. 10 SD, 15 AD, 14 HC | 3 T | FA | TBSS , VBM | 3 mm | 32 | 100 s/mm$^2$ | 0.003 | ILF, UncF, IFOF, SLF, Th, FMi, FMj, Cg, hippocampus | DTI bvFTD vs HC (p<0.05)<br>○ ↓ FA bilaterally in:<br>• IFOF<br>• UncF<br>• ILF and SLF<br>• Ant ThR<br>• FMi and FMj<br>• Cg<br><br>VBM bvFTD vs HC<br>• Rt cerebellum, rt inf TG, rt TP, bilateral para Hpc G, rt hippocampus, bilateral Ag, rt Th, rt and lt insular C, rt and lt OFC, rt mPFC, lt ant Cg, lt TP, lt inferior TG<br><br>FA correlations with autobiographical memory performance<br>• FA +ve correlation with ABM retrieval in the VBM analysis<br>• FA for each WM tracts (p<0.01) show +ve correlations with recent and remote ABM.<br>• <u>Remote ABM retrieval</u> correlated significantly with FA in lt UncF (p=0.001), lt Cg (CC part p=0.008 and hippocampus p=0.002) FMi (p=0.001).<br>• <u>Recent ABM retrieval</u> associated significantly with FA values in FMi (p=0.001) and lt Cg ( hippocampus part p=0.01). | 1. Yes.<br>2. Yes.<br>3. Yes.<br>4. Yes (mean +/- SD provided)<br>5. Yes.<br>6. Yes.<br>7. Yes.<br>8. Yes.<br>9. Yes (Rascovsky reference provided)<br>10. No (El Escorial known)<br>11. Yes.<br>12. Yes.<br>13. Unclear.<br>14. Unclear |

| | | | | | | | | | | | |
|---|---|---|---|---|---|---|---|---|---|---|---|
| Kasper et al (2014) | 72 ALS (49 no cognitive impairment and 23 with cognitive impairment); 65 HC | 3T | FA<br>MD<br>AD<br>RD | Twice refocused spin echo EPI<br><br>TBSS and ROI methods both used | NA | 30 non-collinear directions | 1000 | 0.2 | Whole brain (TBSS);<br>15 ROI's | (ROI methods gave less significant results than TBSS methods, p<0.05, corrected)<br><br>ALS (no CI) vs. HC:<br>• ↓ FA and ↑ RD in bilateral CST, PLIC and bCC<br><br>ALS (CI) vs. HC:<br>• As for the other ALS subjects, but <u>also</u> ↓ FA, ↑ RD and ↑ MD in the ant. ThR, Cg, IFOF, SLF, ILF and UncF (all changes bilateral but lt > rt hemisphere)<br><br>ALS (no CI) vs. ALS (CI):<br>• ↑ RD and ↑ MD in the ant. ThR, Cg, IFOF, SLF, UncF (in CI vs. non-CI patients)<br>• | 1. Yes.<br>2. Yes.<br>3. Yes.<br>4. Yes (mean +/- SD provided)<br>5. Yes.<br>6. Yes.<br>7. Yes.<br>8. Yes.<br>9. Yes (reference provided)<br>10. No (El Escorial known)<br>11. Yes.<br>12. Yes.<br>13. Unclear.<br>14. Unclear (no withdrawals) |
| Kassubek et al (2014) | 111 ALS<br>(A)78 scanned on 1.5 T,<br>(B)33 scanned on 3 T)<br>74 HC<br>(A)52 scanned on 1.5T,<br>(B) 22 on 3 T) | 1.5 T and 3 T | FA, AD, RD | (A) 1.5 T scanner<br><br>(B) 3T<br><br>Tract of interest-based fiber tracking (TFAS analysis vesus ROI-analysis) | 8 mm | (A) 31<br>(B) 49 | 1000 s/mm2 | FA >0.2<br><br>Eignvector = 0,9 | CST, corticorubral (CRT) and Cortico-pontine (CPT), cortico-striatal, prefrontal path | TFAS vs ROI<br>• At 1.5 T:<br> ○ ROI (sensitivity 78%, specificity 69%)<br> ○ TFAS (sensitivity 79%, specificity 71%)<br>• At 3 T<br> ○ ROI (sensitivity 82%, specificity 68%)<br> ○ TFAS (sensitivity 79%, specificity 73%)<br><br>DTI metrics ALS vs HC:<br> ○ Bilateral CST FA=sig<br> ○ RD,AD both significant along CST<br>• Correlation of ALS stages:<br> ○ ALSFRS-R and disease duration correlated significantly with the staging scheme<br> ○ (ALSFRS, p=0.0017)<br> ○ (Disease duration, p=0.0019) | 1. Yes<br>2. Unclear (inclusion criteria clear but no exclusion criteria)<br>3. Yes<br>4. Unclear<br>5. Yes<br>6. Yes<br>7. Yes<br>8. No<br>9. Yes<br>10. Unclear<br>11. Yes<br>12. Yes<br>13. Unclear<br>14. Yes (no withdrawal) |

| | | | | | | | | | | | |
|---|---|---|---|---|---|---|---|---|---|---|---|
| Kim et al. (2014) | 14 ALS<br>16 HC | 3T | FA, | Double SE-EPI<br>Whole brain.<br>Seeding-based tractography | NA | 25 | NA | 0.15 | CC:<br>BA4 (primary motor C)<br>BA1/2/3 (primary sensory C)<br>BA6 (supplementary motor area)<br>BA11/47 (orbitofrontal C)<br>BA44/45 (Broca's area) | ↓ FA in ALS vs HC:<br>○ BA4 ($p=0.003$)<br>○ BA6 ($p=0.01$)<br>○ BA9/46 ($p=0.0005$)<br>○ No sig →BA1/2/3, BA11/47, BA44/45<br>Cortical ROI ↓ FA in ALS vs HC:<br>○ preCG ($p=0.0036$)<br>○ mid.FC ($p=0.016$)<br>○ sup.FC ($p=0.0079$) | 1. Yes<br>2. Yes<br>3. Yes<br>4. Yes<br>5. Yes<br>6. Yes<br>7. Yes<br>8. Yes<br>9. Yes<br>10. No (El Escorial already known)<br>11. Yes<br>12. Yes<br>13. Yes (table with significant and non-significant results presented)<br>14. Unclear |
| Lam et al (2014) | 12 bvFTD,<br>10 PNFA,<br>11 SD,<br>15 HC (2 time-points, 12 months apart) | 3 T | FA, MD, RD, AD | TBSS, VBM | 3 mm | 32 | 1000 s/mm$^2$ | 0.2 | Whole brain | DTI bvFTD vs HC ($p<0.05$):<br>**At base-line**: ↓ FA, ↑ MD, ↑ AD, ↑ RD significant bilateral changes in frontotemporal regions, ant.ThR, ant.Cg, sup ILF, IFOF, UncF, gCC.<br>Changes in MD and RD are most extensive in this group.<br>**At 12 months**: bilateral changes sCC, longitudinal changes were most in FA and RD. rt ant Cg.<br>Changes in MD obvious in Lt SLF.<br>Changes in AD found bilateral ant Cg, SLF and ILF.<br>Overlap between WM and GM changes (after 12 months):<br>GM: Rt Frontotemporal region.<br>WM: bilaterally. | 1. Yes.<br>2. Yes.<br>3. Yes.<br>4. Yes (mean +/- SD provided)<br>5. Yes.<br>6. Yes.<br>7. Yes.<br>8. Yes.<br>9. Yes (references provided)<br>10. No (El Escorial known)<br>11. Yes.<br>12. Yes.<br>13. Unclear.<br>14. Unclear |
| Lu et al (2014) | 8 bvFTD,<br>12 AD,<br>12 HC | 1.5 T | FA, MD, RD, AD | SS-SE<br>ROI | NA | 12 | 800 s/mm$^2$ | NA | Frontal WM, gCC, sCC | DTI group comparisons:<br>• bvFTD showed significant group differences in all DTI metrics in frontal WM and gCC ($p<0.008$).<br>• bvFTD had more WM changes compared to HC and AD in same regions. | 1. Yes.<br>2. Yes.<br>3. Yes.<br>4. Yes (mean +/- SD age at onset and at scan provided)<br>5. Yes. |

| | | | | | | | | | | | |
|---|---|---|---|---|---|---|---|---|---|---|---|
| | | | | | | | | | | • No changes in sCC across all DTI metrics.<br><br>Correlations with behavioral variables:<br>• SEB rating associated with gCC DTI measures; FA (p=0.005), AD (p=0.007), RD (p=0.005) and MD (p=0.0006). Worse WM integrity associated with more symptoms of emotional blunting.<br>• Examining two clinical groups separately showed a correlation between DTI measures and SEB rating in bvFTD. | 6. Yes.<br>7. Yes.<br>8. Yes.<br>9. Yes (Rascovsky reference provided)<br>10. No (El Escorial known)<br>11. Yes.<br>12. Yes.<br>13. Unclear.<br>14. Unclear |
| Mahoney et al (2014) | 23 bvFTD, 18 HC | 3 T | FA, MD, RD, AD | ROI VBM | NA | 64 | 1000 s/mm$^2$ | 0.05 | gCC, sCC, UncF, paraHpc Cg, CST, CP, Fx. | Cross DTI bvFTD vs HC<br>• ↓ FA and ↑ MD (p< 0.02) in bCC, bilateral UncF and rt Cg (para Hpc part).<br>• ↑ AD (p<0.01) in bCC and lt Cg (para Hpc part), Fx (p<0.04) and rt UncF (p<0.03)<br>• ↑ RD (p<0.01) in bCC, bilateral Cg at para Hpc part and lt UncF abd rt UncF (p<0.001).<br><br>Cross DTI MPAT carriers vs HC<br>• ↓ FA rt Cg (p=0.009), ↑ MD in rt UncF (p=0.001)and rt Cg (p=0.01).<br><br>Cross DTI sporadic bvFTD vs HC<br>• ↓ FA lt UncF (p=0.01) and rt Cg (p=0.03).<br>• ↑ MD in lt UncF (p=0.002)<br><br>Cross DTI C9ORF72 carriers vs HC<br>• ↓ FA bilater superior CP (p=0.03)<br><br>Longitudinal DTI bvFTD vs HC<br>• rate of changes in FA and MD were significant is bCC (p=0.003), sCC | 1. Yes.<br>2. Unclear (exclusion criteria not explicitly stated)<br>3. Yes.<br>4. Yes (disease duration introduced as nuisance covariate)<br>5. Yes.<br>6. Yes.<br>7. Yes.<br>8. Yes.<br>9. Yes (Rascovsky reference provided)<br>10. No (El Escorial known)<br>11. Yes.<br>12. Yes.<br>13. Yes.<br>14. Unclear |

<table>
<tr>
<td></td><td></td><td></td><td></td><td></td><td></td><td></td><td></td><td></td><td></td>
<td>(p=0.001), bilateral Cg (p<0.001) and rt UncF (p<0.001) and lt UncF (=0.005).<br>• RD significant changes in subgroups<br>
<table>
<tr><td>RD</td><td>bvFTD</td><td>MAPT</td><td>Sporadic</td><td>C9ORF72</td></tr>
<tr><td>bCC</td><td>0.002</td><td>0.001</td><td>-</td><td>-</td></tr>
<tr><td>sCC</td><td>0.001</td><td>0.01</td><td>0.01</td><td>-</td></tr>
<tr><td>Rt Cg</td><td>0.001</td><td>0.04</td><td>0.002</td><td>-</td></tr>
<tr><td>Lt Cg</td><td>0.001</td><td>0.01</td><td>0.001</td><td>-</td></tr>
<tr><td>Rt Cg Hpc</td><td>0.01</td><td>0.01</td><td>-</td><td>-</td></tr>
<tr><td>Lt Cg Hpc</td><td>0.03</td><td>0.001</td><td>-</td><td>-</td></tr>
<tr><td>Rt UncF</td><td>0.001</td><td>0.001</td><td>0.03</td><td>-</td></tr>
<tr><td>Lt UncF</td><td>0.01</td><td>0.001</td><td>-</td><td>0.02</td></tr>
</table>
• AD significant changes in subgroups<br>
<table>
<tr><td>AD</td><td>bvFTD</td><td>MAPT</td><td>Sporadic</td><td>C9ORF72</td></tr>
<tr><td>gCC</td><td>-</td><td>-</td><td>0.03</td><td>-</td></tr>
<tr><td>bCC</td><td>-</td><td>-</td><td></td><td>0.003</td></tr>
<tr><td>sCC</td><td>0.02</td><td>0.04</td><td>-</td><td></td></tr>
<tr><td>Rt Cg para</td><td>0.001</td><td>0.001</td><td>-</td><td>-</td></tr>
<tr><td>Lt Cg Hpc</td><td>0.005</td><td>0.001</td><td>-</td><td>-</td></tr>
<tr><td>Rt UncF</td><td>0.001</td><td>0.001</td><td>-</td><td>-</td></tr>
<tr><td>Lt UncF</td><td>0.009</td><td>0.001</td><td>-</td><td>0.03</td></tr>
<tr><td>sCP</td><td>0.008</td><td>0.03</td><td>-</td><td>0.003</td></tr>
</table>
DTI metrics sensitivity and specificity<br>• Cross sectionally: RD best then MD followed by FA and AD<br>• Longitudinaly were best achieved by FA, followed by MD and RD then AD.</td>
<td></td>
</tr>
<tr>
<td>Menke et al 2014</td>
<td>60 ALS<br>36 HC</td>
<td>3T</td>
<td>FA, MD, RD, AD</td>
<td>MPRAGE, VBM, TBSS and ROI</td>
<td>3mm</td>
<td>60</td>
<td>1000 s/mm2</td>
<td>CC at 45<br>SLF at 10</td>
<td>CC, CST, SLF, FL and pre-motor cortex</td>
<td>In ALS gp at T0:<br>1. VBM ↓ GM<br>○ lt. primary MC and lt. FL.<br>○ Th, bilateral caudate head<br>2. DTI:<br>○ ↑AD, MD in CC<br>○ ↑AD in ltCST<br><br>UMN scores:<br>1. TBSS:<br>○ ↓FA, ↑AD in CST<br>○ ↑MD, ↑RD in CST,SLF, CC<br>2. ROI → ↓FA in rt.SLF</td>
<td>1. Yes<br>2. Yes<br>3. Yes<br>4. Yes<br>5. Yes<br>6. Yes<br>7. Yes<br>8. Yes<br>9. Yes<br>10. No (El Escorial already known)<br>11. Unclear<br>12. Yes</td>
</tr>
</table>

| | | | | | | | | | | | |
|---|---|---|---|---|---|---|---|---|---|---|---|
| | | | | | | | | | | 3. VBM → No sig<br><br>ALSFRS:<br>○ TBSS, VBM→ no sig<br>○ **ROI**→ RD –ve correlation in lt.SLF<br><br>progression rate:<br>1. TBSS, VBM→ no sig<br>2. ROI→<br>○ ↑RD , ↓FA in rt.SLF, CST<br>○ ↑AD in CC<br>3. **VBM ↓ GM** –ve correlation in lt. primary MC<br><br>ACE:<br>○ TBSS, VBM →No sig<br>○ **ROI :** verbal fluency and AD →+ve correlation<br>○ **VBM:** verbal fluency and Broca's area, dorsolateral prefrontal cortex→+ve correlation | 13. Yes (supplementary material available online)<br>14. Unclear |
| Prokscha et al (2014) | 12 HC; 13 ALS | 1.5 T | FA | Single-shot EPI Manual and atlas ROI approaches (with or without TBSS) | Not stated | 12 | 1000 s/mm2 | Not stated | CST | • ↓FA in bilateral CST in ALS (most significant using atlas ROI with TBSS approach; p = 0.0002)<br><br>• ROC analysis showed best test performance was using atlas ROI with TBSS approach (AUC = 0.936, sensitivity 100%, specificity 91.67%) | 1. Yes<br>2. Yes<br>3. Yes<br>4. Yes<br>5. Yes<br>6. Yes<br>7. Yes<br>8. Yes<br>9. Yes<br>10. No (El Escorial already known)<br>11. Yes<br>12. Yes<br>13. Unclear<br>14. Yes (reasons for withdrawals/ post-recruitment exclusions identified) |

| | | | | | | | | | | | |
|---|---|---|---|---|---|---|---|---|---|---|---|
| Romano et al (2014) | 14 ALS; 14 HC | 1.5 T | FA<br>MD<br>AD (PD)<br>RD | SS-SE EPI (DTI performed to define the tracts as part of waveguide elastography (WGE) protocol) | NA | 12 non-collinear directions | 1000 | NA | CST | ALS vs. HC:<br>• In the CST's, there was significant ↓ FA (p = 0.011), ↑ MD (p = 0.023) and ↑ RD (0.007)<br><br>No significant difference in AD | 1. Yes<br>2. Unclear (exclusion criteria not specified)<br>3. Yes.<br>4. Unclear.<br>5. Yes.<br>6. Yes.<br>7. Yes.<br>8. Yes.<br>9. Yes (reference provided)<br>10. No (El Escorial known)<br>11. Yes.<br>12. Yes.<br>13. Unclear.<br>14. Unclear (no withdrawals) |
| Sarica et al (2014) | 14 HC; 14 ALS | 3T | FA<br>MD<br>RD<br>AD | Spin-echo EPI Tractography (TRACULA) TBSS (to corroborate TRACULA results) | Not stated | 27 | 1000 s/mm2 | Not stated | CST, SLF (parietal and temporal), anterior thalamic radiation (ATH), Unc.F, supracallosal bundle, ILF, CC-forceps major (Fj) | • Tractography (ALS vs. HC, p<0.05, corrected for multiple comparisons):<br> ○ Lt. cingulum-cingulate gyrus (supracallosal) bundle (RD, ↑MD)<br> ○ Rt. CST (↓FA, ↑RD, ↑MD)<br> ○ Rt. supracallosal bundle (↑MD, ↑AD)<br> ○ Lt.ATR (↓FA, ↑RD, ↑MD) and rt Cg (↑AD and ↑MD) uncorrectep<0.05.<br>• TBSS (ALS vs. HC, p<0.05, TFCE corrected):<br> ○ Bilateral CST (FA, MD, RD)<br> ○ Bilateral SLF (FA, MD)<br> ○ Bilateral anterior thalamic radiation (FA, MD, RD)<br> ○ Bilateral Unc.F (MD)<br> ○ Bilateral CR (FA, MD, RD)<br> ○ Cg (FA, MD, RD)<br> ○ Bilateral ILF (MD, RD)<br> ○ CC (FA, MD, RD)<br> ○ No significant changes in AD<br>• Correlations with clinical measure: | 1. Yes<br>2. Yes<br>3. Yes<br>4. Yes (except one patient DD = 16 yrs with minimal disability)<br>5. Yes<br>6. Yes<br>7. Yes<br>8. Yes<br>9. Yes<br>10. No (El Escorial already known)<br>11. Yes<br>12. Yes<br>13. Yes (negative results also reported in table)<br>14. Unclear (no evidence of withdrawals) |

| | | | | | | | | | | | |
|---|---|---|---|---|---|---|---|---|---|---|---|
| | | | | | | | | | | ○ Rt. CST (RD and MD) correlated negatively with ALSFRS-R (p=0.0009)<br>○ Rt. CST (FA) showed a trend towards significant with ALSFRS-R (p=0.07) | |
| Tovar-Moll et al (2014) | 20 bvFTD, 19 CBS, 15 HC | 3 T | FA, MD | Voxel wise whole brain analysis<br>ROI | Smoothing | Directions | 1000 s/mm$^2$ | FA Thresh | Whole brain<br><br>ROIs: CC (gCC and sCC), IC, CR, Cg, UncF, medial forebrain bundle (MFB). | DTI ROI results (p<0.05):<br>○ Patients vs HC:<br>• ↓ FA and ↑ MD in all tracts except IC and sCC.<br>○ bvFTD vs HC<br>• Showed more changes in the Cg G, UncF in frontal and temporal parts.<br><br>DTI VWA results (p<0.05):<br>○ bvFTD vs HC<br>• Abnormalities in CC, AC, CR, UncF, IFOF, ILF, SFOF, SLF, Cg and MFB.<br>• Ant limb of IC.<br>○ bvFTD vs CBS<br>• WM damage restricted to FMi and gCC, UncF, MFB and rostral SLF.<br>• Rt hemisphere was damaged more than the lt in gCC, UncF and rostral SLF.<br><br>Correlations with clinical ratings<br>In bvFTD:<br>○ Posterior Cg correlated with:<br>• NPI-apathy subscores (MD, p<0.01)<br>• Mattis total (MD and FA, p<0.05)<br>○ UncF:<br>• NPI-aberrant motor behavior subscore with frontal part (MD and FA, p<0.05) and temporal part (FA, p<0.05)<br>• | 1. Yes.<br>2. Yes.<br>3. Yes.<br>4. Yes (mean +/- SD provided)<br>5. Yes.<br>6. Yes.<br>7. Yes.<br>8. Yes.<br>9. Yes (Neary & McKhann references provided)<br>10. No (El Escorial known)<br>11. Yes.<br>12. Yes.<br>13. Unclear.<br>14. Unclear |

| | | | | | | | | | | | |
|---|---|---|---|---|---|---|---|---|---|---|---|
| Woo et al (2014) | 34 ALS<br>13 HC | 3T | FA, MD | SS-SE<br>GRAPPA<br>Deterministic tractography | | 30 | 1000 s/mm3 | 0.2 inner 0.75 | CST, forceps major (Fj) | • Between ALS and HC:<br>o CST FA (p=0.01) and MD (p=0.003)<br>o Linear relations with UMN score and ALSFRS-R.<br>o No association with disease duration<br>• Regression results:<br>o MD and FA in CST was significant (p=0.02)<br>o MD in Fj =no sig<br>o UMN score (MD, p=0.005) (FA, p=0.003) and age (MD, p=0.03) both were significant predictor but not age and FA.<br>o ALSFRS-R and disease duration = no sig<br>o El Escorial, handedness and sex = no sig | 1. Yes<br>2. Yes<br>3. Yes<br>4. Yes<br>5. Yes<br>6. Yes<br>7. –<br>8. yes<br>9. yes<br>10. yes<br>11. yes<br>12. Unclear<br>13. Unclear<br>14. Unclear |
| Zhang et al (2014) | 20 ALS<br>21 HC | 3T | FA, RD, AD, | SE-EPI<br>TBSS AND PDT | NA | 64 | 1000 s/mm2 | 2 | Superior CR, CP, PLIC, PreCG, body CC, CST, SLF, CF | • VWA: ↓ FA in ALS vs HC<br>o Bilateral Sup.CR<br>o Caudal CST (rt.CP), Lt. PLIC.<br>o Body CC.<br>o Rt. preCG<br>• PDT: ↓ FA and ↑ RD in ALS vs HC<br>o Bilateral CST to preCG<br>o CC fibers (bilateral SMA and preCG)<br>o Rt.AF (SLF) – different result<br>• Cortical thinning:<br>o Lt.OFC<br>o Rt.SMA<br>o Bilateral preCG<br>o Dorsal preMC.<br>o Sup.PL<br>o Lt. mid. OG<br>o SMG<br>o Rt. Prefrontal regions<br>o Lt.OFC | 1. Yes<br>2. Yes<br>3. Yes<br>4. Yes<br>5. Yes<br>6. Yes<br>7. Yes<br>8. Yes<br>9. Yes<br>10. No (El Escorial already known)<br>11. Yes<br>12. Yes<br>13. Unclear<br>14. Unclear |
| Agosta et al (2015) | 21 ALS, 14 ALS-plus, 14 bvFTD, 12 SD, 11 | 3T | FA<br>MD | Pulsed-gradient SE EPI<br><br>TBSS – WM voxelwise analysis | 8 mm FWHM | 32 non-collinear directions | 1000 | 0.2 | Whole brain | • WM changes more widespread than GM (all groups)<br>• Results $p < 0.05$, FWE corrected<br>• Supporting figures on journal website | 1. Yes.<br>2. Yes.<br>3. Yes.<br>4. Unclear.<br>5. Yes.<br>6. Yes. |

| | | | | | | | | | | | | |
|---|---|---|---|---|---|---|---|---|---|---|---|---|
| | PNFA, 28 HC | | | | | | | | | | ALS vs HC:<br>• ↑ MD and ↓ FA in bilateral CR and posterior bCC.<br><br>ALS-plus/bvFTD/PNFA vs. HC:<br>• ↑ MD and ↓ FA in the CC, orbitofrontal WM, frontoparietal WM, occipital WM, IC, BS, temporal WM (all bilateral)<br><br>SD vs. HC:<br>• ↑ MD in the lt (> rt) IC and External capsule (ExC) , bCC, orbitofrontal, frontal, ant. temporal and inferior parietal WM (sparing of occipital lobes and BS)<br>• ↓ FA in bCC and gCC, lt SLF, Cg, ExC and IC, ant. and middle temporal WM<br><br>Between group ALS and FTD comparisons also performed | 7. Yes.<br>8. Yes.<br>9. Yes (references provided)<br>10. No (El Escorial and FTD criteria known)<br>11. Yes.<br>12. Yes.<br>13. Unclear.<br>14. Unclear<br><br>(no withdrawals) |
| Bae et al (2015) | 25 ALS,17 bvFTD, 37 HC | 3 T | NA | ROI<br>VBM | NA | 32 | 1000 s/mm$^2$ | NA | WM Regions in Th, Cerebellum,motor cortex (pre CG, post CG, and supplementary motor area), BS and striatum (putamen, caudate, globus pallidus) | VBM and DTI:<br>ALS vs HC (p<0.05):<br>○ Minor changes is the<br> • Motor cortex<br> • Cerebellum<br> • Th<br> • BS<br>○ WM showed greater changes is these regions including CST<br><br>bvFTD vd HC (p<0.05):<br>○ Sever degeneration across<br> • Motor system<br> • Cerebellum<br>○ WM that connect these regions is severely affected.<br><br>ALS vs bvFTD (p<0.05):<br>○ Minimal GM atrophy in the cerebellum.<br>○ WM changes in lt. Th in ALS.<br>○ While bvFTD patients showed more atrophy in: | 1. Yes.<br>2. Yes (El Escorial and Rascovsky)<br>3. Yes.<br>4. Yes (mean +/- SD provided)<br>5. Yes.<br>6. Yes.<br>7. Yes.<br>8. Yes.<br>9. Yes (reference provided)<br>10. No (El Escorial known)<br>11. Yes.<br>12. Yes.<br>13. Yes.<br>14. Unclear. |

| | | | | | | | | | | | |
|---|---|---|---|---|---|---|---|---|---|---|---|
| | | | | | | | | | | • Motor cortex<br>• Cerebellum<br>• Striatal<br>• Th<br><br>Changes in WM includes these regions found in VBM | |
| Daianu et al (2015) | 20 bvFTD, 23 early-onset AD, 33 HC | 1.5 T | FA,MD, RD, AD | SS-SE DWI<br><br>autoMATE (automated multi-atlas tract extraction) Tractography | NA | 30 | 1000 s/mm$^2$ | 0.2 | **21 tracts:** Th, CC, Cg, CST, IFOF, ILF, SLF, UncF, paraHcp | bvFTD vs HC:<br><br>• MD and RD detected most changes.<br>• Frontal and temporal tracts are most impacted.<br>• All 21 tracts were severly affected with ↑ MD (p=0.035).<br>• ↓ FA (p=0.024) with ↑ AD (p=0.019), RD (p=0.034) and MD were significant (above 60%) in rt. Ant ThR. And bilateral UncF.<br>• Commissural fibers: most significant is the frontal fibers in more than 99%. gCC shows late myelination<br>• Long association fibers: more than 99% in bilateral UncF, lt. SLF and bilateral IFOF.<br>• Limbic system: dorsal Cg and ant. ThR<br><br>Motor system: CST was the least affected tract. | 1. Yes.<br>2. Yes (Rascovsky)<br>3. Yes.<br>4. Unclear.<br>5. Yes.<br>6. Yes.<br>7. Yes.<br>8. Yes.<br>9. Yes (reference provided)<br>10. No (El Escorial known)<br>11. Yes.<br>12. Yes.<br>13. Unclear.<br>14. Unclear |
| Downey et al (2015) | 29 bvFTD, 15 svPPA, 37 HC | 3 T | Trace diffusivity (TR), FA, MD, RD, AD | SS-SE<br><br>WM TBSS<br><br>GM: VBM using DARTEL toolboxes of SPM8 | 6 mm | 64 | 1000 s/mm$^2$ | NA | Whole brain | Brain maps bvFTD vs HC<br>○ GM: bi hemisohereic atripgy in:<br> • Ant TL, mTL structures, insular, prefrontal and orbitofrontal cortices.<br>○ WM (p<0.05):<br> • Changes were dorsal and ventral mostly in Cg, bi UncF, CC<br> • Less posteriorly in parieto-occipital ILF.<br><br>Correlations with social cognition performance (p<0.05, corrected) | 1. Yes.<br>2. Yes (Rascovsky and Gomo-Tempini references provided)<br>3. Yes.<br>4. Yes (mean +/- SD provided)<br>5. Yes.<br>6. Yes.<br>7. Yes.<br>8. Yes.<br>9. Yes (references provided)<br>10. No (El Escorial known) |

| | | | | | | | | | | | |
|---|---|---|---|---|---|---|---|---|---|---|---|
| | | | | | | | | | | • -ve correlation betweem TASIT emotion identification score and DTI metrics (AD,TR and RD).<br>• +ve correlated with FA in dorsal and ventral commissural WM bilaterally.<br>• Frontal subcortical projection pathways (rt ant ThR) and Fx<br>• Within bvFTD group, emotion identification impairment was associated with WM alteration in CC and Fx.<br>• -ve correlation between TASIT total sarcasm with AD, RD and TR in the rt temporal WM, and inferior frontal WM<br>• +ve correlation in sarcasm identification scores with rt side of WM but bilateral in Temporal inferior FWM, rt UncF and rt ant. ThR. | 11. Yes.<br>12. Yes.<br>13. Unclear.<br>14. Unclear |
| Hubers et al (2015) | Subset of 30 ALS patients; 18 HC | 1.5 T | FA | Voxelwise statistical comparison (WBSS) with manual ROI method | 8 mm FWHM | 31 | 1000 | 0.2 | CST | ALS vs. HC:<br>• ROI in the bilateral lower CST in both hemispheres<br><br>Significant ↓ FA in ALS compared with HC ($p < 0.01$) | 1. Yes.<br>2. Yes.<br>3. Yes.<br>4. Yes (mean +/- SD provided)<br>5. Yes.<br>6. Yes.<br>7. Yes.<br>8. Yes.<br>9. Yes (reference provided)<br>10. No (El Escorial known)<br>11. Yes.<br>12. Yes.<br>13. Unclear.<br>14. Yes (not all OCT patients consented to the MRI component) |

| | | | | | | | | | | | |
|---|---|---|---|---|---|---|---|---|---|---|---|
| Sheelakumari et al (2015) | 17 ALS, 15 HC | 1.5 T | FA<br>MD | SS-SE EPI<br><br>ROI-based analysis | NA | 30 non-collinear directions | 1000 | NA | CST (ROI's in pons at level of CP and medullary pyramids) | ALS vs. HC:<br>• PONS: ALS subjects had ↓ FA ($p = 0.001$) and ↑ MD ($p = 0.003$)<br><br>MEDULLA: ALS subjects had ↑ MD ($p = 0.011$); FA not significant when corrected. | 1. Yes.<br>2. Yes.<br>3. Yes.<br>4. Yes (online supplementary table)<br>5. Yes.<br>6. Yes.<br>7. Yes.<br>8. Yes.<br>9. Yes (reference provided)<br>10. No (El Escorial known)<br>11. Yes.<br>12. Yes.<br>13. Unclear.<br>14. Unclear (no withdrawals, retrospective cohort) |
| Steinbach et al (2015) | 16 ALS, 16 HC | 3T | FA<br>MD | Spin echo EPI<br><br>Probabilistic tractography → structural connectivity indices (CI) calculated | NA | 12 non-collinear directions | 1000 | 0.2 | 1. Intracranial CST (pons to PMC)<br>2. Visual cortex to medial temporal (ERC, PRC, PHC) | ALS vs. HC:<br>• ↓ CI in the CST's ($p = 0.03$), negative correlation with disease duration<br><br>↓ CI in the VC-PRC tract ($p = 0.02$), no significant effect in the VC-ERC or VC-PHC tracts | 1. Yes.<br>2. Yes.<br>3. Yes.<br>4. Yes<br>5. Yes.<br>6. Yes.<br>7. Yes.<br>8. Yes.<br>9. Yes (reference provided)<br>10. No (El Escorial known)<br>11. Yes.<br>12. Yes.<br>13. Unclear.<br>14. Unclear |
| Tang et al (2015) | 69 ALS, 23 HC | 3T | FA<br>ADC | SS-SE EPI | NA | 23 | 1000 | Different thresholds used (ROC) | 18 regions examined | • Data from rt and lt sides averaged<br><br><u>ALS vs. HC</u>: ($p < 0.05$) | 1. Yes.<br>2. Yes.<br>3. Yes.<br>4. Unclear.<br>5. Yes.<br>6. Yes.<br>7. Yes. |

| | | | | | | | | | | | |
|---|---|---|---|---|---|---|---|---|---|---|---|
| | | | | | | | | | | • ↓ FA in centrum semiovale, deep frontal WM, deep parietal WM, CR, PLIC, gCC of CC, sCC, CP.<br><br>↑ ADC in centrum semiovale, deep frontal WM, deep parietal WM | 8. Yes.<br>9. Yes (reference provided)<br>10. No (El Escorial known)<br>11. Yes.<br>12. Yes.<br>13. Unclear<br>14. Unclear. |
| Zimmerman-Moreno et al (2015) | 23 ALS, 18 HC | 3T | FA<br>MD<br>RD<br>AD | Novel method (FBC), compared with TBSS method | NA | 15 (19 for 3 control subjects) | 1000 | 0.2 | Whole brain | ALS vs. HC (TBSS method):<br>• Significant ($p \leq 0.05$ corr) ↓ FA and ↑ RD in the CST and bCC | 1. Yes.<br>2. Yes.<br>3. Yes.<br>4. Unclear.<br>5. Yes (no El Escorial criteria/reference provided, but same patients scored in Ben Bashat et al (2011))<br>6. Yes.<br>7. Yes.<br>8. Yes.<br>9. Yes<br>10. No (El Escorial provided in previous paper, as above)<br>11. Yes.<br>12. Yes.<br>13. Unclear.<br>14. Unclear |
| Vora et al (2016) | 21 ALS, 13 HC | 1.5 T | FA<br>ADC<br>MD | Single shot diffusion weighted EPI<br><br>ROI-based methods | NA | 30 | NA | NA | ROI's at PMC WM, gCC and sCC, PLIC, medullary pyramid | ALS vs. HC:<br>• ↓ FA and ↑ MD in bilateral PMC WM, PLIC, gCC and sCC ($p < 0.05$)<br>• ↓ FA lt medullary pyramid ($p = 0.007$)<br><br>CC and medullary pyramid changes were <u>not</u> significant when looking at the "possible ALS" | 1. Yes.<br>2. Yes (divided into definite, probable and possible ALS)<br>3. Yes<br>4. Yes<br>5. Yes<br>6. Yes<br>7. Yes<br>8. Unclear (some details not included)<br>9. Yes<br>10. No (El Escorial known) |

| | | | | | | | | | | | |
|---|---|---|---|---|---|---|---|---|---|---|---|
| | | | | | | | | | | | 11. Yes<br>12. Yes<br>13. Unclear<br>14. Unclear |

#### Supplementary Table 2: FA correlations in ALS

| Disease | Tracts | DTI measure | Correlation | ROI | Disease measure |
|---|---|---|---|---|---|
| ALS | Projection fibers | FA | -ve | CST | UMN scores |
| | | | -ve | | DD |
| | | | +ve | | CMCT |
| | | | | | Cortical_brain stem conduction time |
| | | | | | Trail making test scores |
| | | | | | Stroop test scores |
| | | | | | Letter fluency |
| | | | -ve | | FrSBe |
| | | | -ve (weaker) | Rt CST | ALSFRS-R |
| | | | +ve | Rt. UncF | Sarcasm identification scores |
| | | | | Bilateral IFOF ILF | Disease progression |
| | | | +ve | Bilateral IFOF ILF | Trail making |
| | | | +ve | Lt IFOF ILF | Stroop test scores |
| | | | -ve | SLF | • FBI A, FBI B and FBI AB<br>• Personal neglect (eg, lack of personal hygiene, disorganization in planning and organizing complex activity, impulsivity or poor judgment and utilization behavior |
| | | | -ve | SLF | FrSBe |
| | | | -ve | SLF | • UMN scores<br>• Trail-making Test B scoring |

| | | | | | |
|---|---|---|---|---|---|
| | | | -ve | Rt ILF | Cumulative scores of single emotions |
| | | | -ve | IFOF | Emotional recognition |
| | Commissural F | ↓ FA | Weaker | | ALSFRS-R |
| | | FA | | | Trail making test scores |
| | | | | | Stroop test |
| | | | +ve | Dorsal and ventral CC | social cognition performance |
| | Limbic | FA | | Fornix | Verbal learning |
| | | | | Fornix | Memory test scores |
| | | | | Para Hpc<br>Lt Th | Increased Apathy scores on FrSBe |
| | | | +ve | Rt ant ThR | Sarcasm identification scores |
| | Whole Brain | FA | | Lt hemisphere | VBM |

#### Supplementary Table 3: MD correlations in ALS

| Disease | Tracts | DTI measure | Correlation | ROI | Disease measure |
|---|---|---|---|---|---|
| ALS | Limbic | MD | | Fornix | Verbal learning |
| | | | | | Memory test scores |
| | | | -ve | Th | ALSFRS-R |
| | | | +ve | | DD |
| | Commissural | MD | | CC | Trail making test scores |
| | | | | CC | Stroop test |
| | Projection F | ↑ MD | | CST | ↑ UMN scores |
| | | | -ve | Rt CST | ALSFRS-R |

**Supplementary Table 4: radial diffusivity and correlations in ALS**

| Disease | Tracts | DTI measure | Correlation | ROI | Disease measure |
|---|---|---|---|---|---|
| ALS | Commissural | ↑ $D_{radial}$ | | CC | Clinical scores |
| | Projection | ↑ $D_{radial}$ | using ROI | CST | ↑ UMN scores |
| | | | | | ↓ CMCT |
| | | | | | ↓ Cortical-brain stem conduction time |
| | | | | | ↓ Trail making test scores |
| | | | | | ↓ Stroop test scores |
| | | | | | ↑ Disease progression |
| | | | -ve | Rt CST | ALSFRS-R |
| | Association Fibers | | -ve | Rt SLF | ALSFRS-R |
| | | | +ve | Rt SLF | Disease progression |
| | Whole brain | | Lt hemisphere | | VBM |

**Supplementary Table 5: Axial diffusivity and correlations in ALS studies**

| Disease | Tracts | DTI measure | Correlation | ROI | Disease measure |
|---|---|---|---|---|---|
| ALS | Projection | ↑ $D_{axial}$ | | CST* | ↑ UMN scores |
| | | | | CST* | ↑ DD |

**Supplementary Table 6: FA correlations in FTD studies.**

| Disease | Tracts | DTI measure | Correlation | ROI | Disease measure |
|---|---|---|---|---|---|
| FTD | Projection F | ↓ FA | +ve | | Cortical thickness |
| | Association T | | | SLF IFOF ILF UncF | Cortical thickness |
| | | FA and MD | | UncF | NPI-aberrant motor behavior subscore |
| | | FA | +ve | ILF, IFOF,UncF, SLF | Recent and remote autobiographical memory performance (ABM) |
| | Limbic | FA | | Post Cg | Mattis total |
| | | MD | | Post Cg | NPI-apathy subscores |
| | | | | | Mattis total |
| | | FA,MD,RD,AD | -ve | gCC | SEB rating |
| | Whole Brain | | -ve | | TASIT emotion identification score |
| | Whole brain | | Lt hemisphere | | VBM |

The staging system proposed by Roche et al. for ALS is as following (211):

Stage 1: symptom onset;

Stage 2A: diagnosis;

Stage 2B: involvement of a second region;

Stage 3: involvement of a third region;

Stage 4A: need for gastrostomy;

Stage 4B: need for respiratory support
